# A general theory for infectious disease dynamics

**DOI:** 10.1101/2022.09.12.22278744

**Authors:** Giuseppe Carbone, Ilario De Vincenzo

## Abstract

We present a general theory of infection spreading, which directly follows from conservation laws and takes as inputs the probability density functions of latent times. The derivation of the theory substantially differs from Kermack and McKendrick (1927) argument, which instead was based on the concept of removal rates. We demonstrate the formal equivalence of the two approaches, but our theory provides a clear interpretation of the kernels of the integro-differential equations governing the infection spreading in terms of survival function of the latent times distributions. This aspect was never captured before. Real distributions of latent times can be, then, employed, thus overcoming the limitations of standard SIR, SEIR and other similar models, which implicitly make use of exponential or exponential-related distributions. SIR and SEIR-type models are, in fact, a subclass of the theory here presented. We show that beside the infection rate *ν*, the joint probability density function *p*_EI_ (*τ, τ*_1_) of latent times in the exposed and infectious compartments governs the infection spreading. Assuming that the number of infected individuals is negligible compare to the entire population, we were able to study the stability of the dynamical system and provide the general solution of equations in terms characteristic functions of the probability distribution of latent times. We present asymptotic solutions for the case *R*_0_ = 1 and demonstrate that the moments of the latent times distribution govern the rate of disease spreading in this case. The present theory is employed to simulate the diffusion of COVID-19 infection in Italy during the first 120 days. The estimated value of the basic reproduction number is *R*_0_ ≈ 3.5, in very good agreement with existing data.

## Introduction

Mathematical modeling has proven to be an extremely powerful tool to explain and predict the course of an epidemic, by helping to plan effective policies to control the infection spreading (*1–8*).

The most common framework for modeling infectious disease dynamics involves dividing the population into compartments and using ordinary differential equations (ODEs) to specify flows between them. For diseases conferring permanent immunity, the SIR (susceptible - infectious - recovered) model is usually employed (*1,9,10*). A simple enhancement of the SIR model is SEIR one (susceptible - exposed - infectious - recovered), where the compartment of exposed (i.e. infected) individuals but not yet infectious is introduced. These models assume that the time an individual spends in each compartment is exponentially distributed (*2–5*). While such an assumption could sound reasonable in some circumstances, it is often unrealistic (*11–15*), and usually leads to an underestimation of the basic reproduction number and to overoptimistic predictions about the efficacy of epidemic control measures (*16*). The assumption of exponentially distributed latent times, may also overestimate the numbers of individuals whose infection period is much shorter or longer than its average (*13, 14*). Several investigations have, indeed, revealed that the shape of the probability density function of latent times may have a huge impact on the predicted infection dynamics (*14,16–30*). Aware of these limits, we present a theory of infection spread, where the transition from the susceptible compartment to the exposed one is still governed by a reaction-like equation, but the flow of individuals to any other subsequent compartment (e.g., from exposed to infectious or from infectious to removed) is rather governed by integral equations that express the fact that individuals, who enter a certain compartment at a given time will move to the subsequent one after a generic stochastically distributed latent time. The present theory does not pose any restriction on the functional form of the latent times probability density functions. Thus, real distributions, obtained by tracing real data, can be easily utilized. The present theory is employed to describe the evolution of COVID-19 infection (*31–35*) in Italy during the first 120 day.

## 1 The Model

We assume homogenous mixing of the population. Accordingly, the effect of the topological structure of the individuals contact network is not considered in this study. However, the theory can be easily generalized to the case of disease spreading on networks or in the case of heterogeneous mixing (*36–38*).

Our model consists of six compartments: (i) susceptible, (ii) exposed, (iii) infectious, (iv) removed, (v) healed, (vi) deceased individuals as shown in Fig. 1. Let be *N* the total and constant number of individuals in the population, *s* (*t*) the number of susceptible individuals, *e* (*t*) the number of exposed individual, *i* (*t*) the number of infectious individuals, *r* (*t*) the number of removed (isolated) individuals, *h* (*t*) the number of recovered (healed) individuals, and *d* (*t*) the number of deceased individuals. We also define *n* (*t*) = *N* − *s* (*t*) the cumulative number of cases, i.e. the total number of individual that have been infected. Mass conservation requires *s* (*t*) + *e* (*t*) + *i* (*t*) + *r* (*t*) + *h* (*t*) + *d* (*t*) = *N*, i.e.

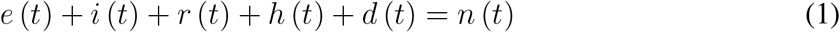

We assume that all the above defined quantities are zero for negative times, i.e. we assume that *n* = *e* = *i* = *r* = *h* = *d* = 0 for *t <* 0. Moreover at *t* = 0 only *n* (*t*), *s* (*t*) and *e* (*t*) undergoes a step change so that *n* (0^+^) = *e* (0^+^) = *n*_0_ and *s* (0^+^) = *s*_0_ = *N* − *n*_0_. By defining the Heaviside step function ℋ (*t*) as ℋ (*t* ≤ 0) = 0 and ℋ (*t >* 0) = 1, we can write that the time derivative 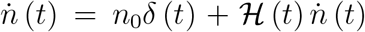, and analogously 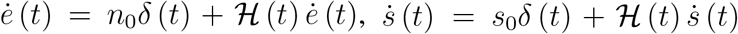, where *δ* (*t*) is the Dirac delta function. Now we assume that transition from the susceptible compartment to the exposed one is governed by the standard reaction-like equation

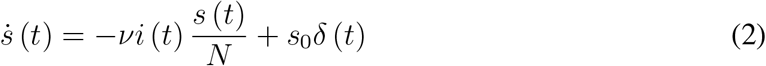

Note that the forcing term *s*_0_*δ* (*t*) is included to account for the condition *s* (0^+^) = *s*_0_. In fact integrating both hand sides from *t* = 0^−^ to *t* = 0^+^ one gets *s* (0^+^) − *s* (0^−^) = *s*_0_ and recalling that *s* (0^−^) = 0 we retrieve the initial condition *s* (0^+^) = *s*_0_. In Eq. (2) the infection rate *ν* is the average number of contacts per person per unit time, multiplied by the probability of disease transmission in a single contact between a susceptible and an infectious individual. Here we assume, since the beginning, that the infection rate of each individual does not depend on the time when the he/she got infected. However the extension to this case is straightforward. Incidentally we notice that in general Kermack and McKendrick (*9*) model this assumption was enforced at the beginning of their argument. Eq. (2) can be rephrased as

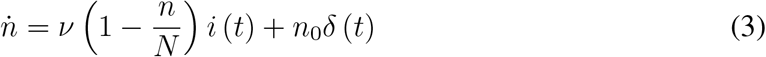

Now consider a single individual that has been infected at time *t*. Such an individual will become infectious after a certain randomly distributed latent period *τ*_*k*_ ≥ 0 and therefore will move from the exposed to the infectious compartment at time *t* + *τ*_*k*_. Let us call *δm*_*k*_ (*t*) the number of individuals in the time interval *δt*, having latent time *τ*_*k*_, which move out from the exposed compartment at time *t*. Of course *δm*_*k*_ (*t*) equals the number of individuals *δn*_*k*_ (*t* − *τ*_*k*_) that have been infected (during the same time step *δt*) at time *t* − *τ*_*k*_. Hence, the increase *δe* (*t*) of individuals in the exposed compartment at time *t* is just given by the number *δn* (*t*) of newly infected individuals at time *t* diminished by the number of individuals *δm* (*t*) = ∑ _*k*_ *δm*_*k*_ (*t*) = ∑ _*k*_ *δn*_*k*_ (*t* − *τ*_*k*_), i.e.

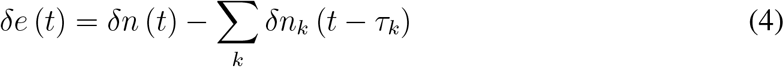

or dividing by the infinitesimal time step *δt*

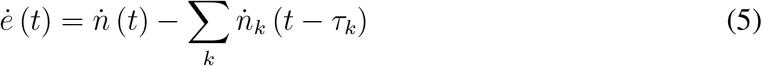

Recalling that the latent time *τ* individuals spend in the exposed compartment is a continuous stochastic variable, one can write that the fraction of individuals that have latent time ranging in the interval *τ*_*k*_ ≤ *τ < τ*_*k*_ + Δ*τ* is

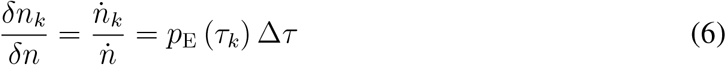

where *p*_E_ (*τ*) is the density probability function of latent times *τ >* 0. We assume that *p*_E_ (*τ*) is independent of time *t*. This yields

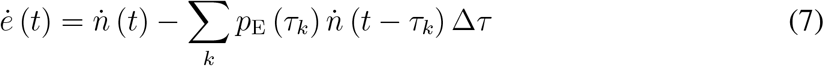

and by letting Δ*τ* → 0 we get

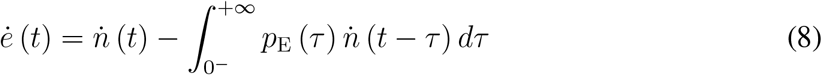

Eq. (8) can be easily *t*-integrated to get

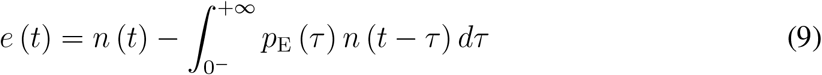

Observe that Eq. (9) is Volterra equation. A similar argument applies to the number *i* (*t*) of individuals in the infectious compartment (see Sec. A) to give

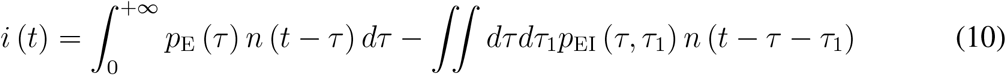

where *p*_EI_ (*τ, τ*_1_) is the joint probability density function of the latent times *τ* and *τ*_1_ in the exposed and infectious compartments respectively (*τ*_1_ is the time the single individual spends into the infectious compartment). Note that to simplify the notation we have used the symbol ∫ in place of 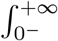. Consider that the lower limit of integration 0^−^can be replaced with −∞ since the probability density functions must vanish when at least one of the arguments is negative.

**Figure 1:**
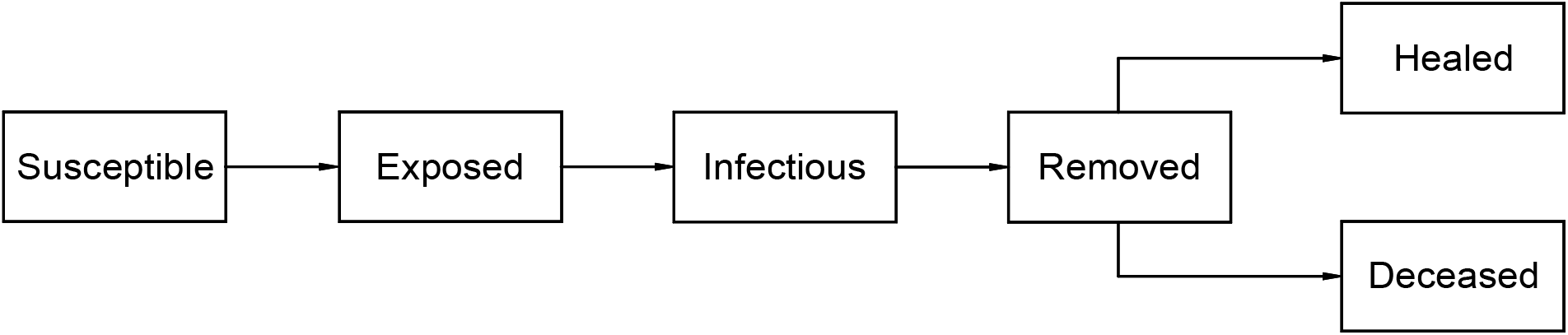
The compartments of the proposed model.

Using the same arguments for the case of individuals in the removed compartment, and defining the latent time *τ*_2_ as the time a removed individual takes to recover and *τ*_3_ the latent time a removed individual takes to enter the deceased compartment we get

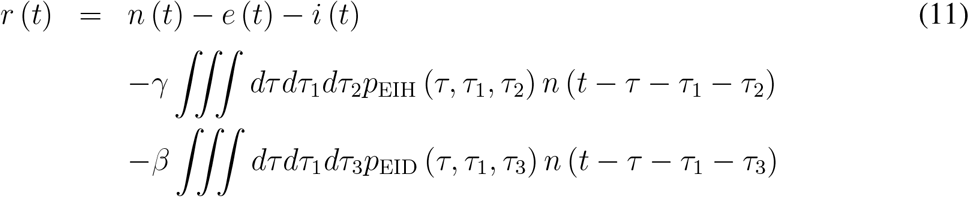

and

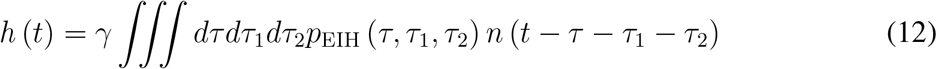

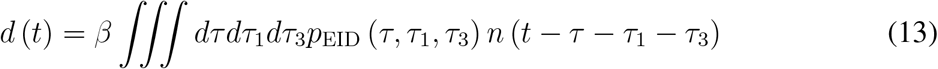

where *p*_EIH_ (*τ, τ*_1_, *τ*_2_) is the joint probability density function of latent times *τ, τ*_1_ and *τ*_2_, and *p*_EID_ (*τ, τ*_1_, *τ*_3_) is the joint probability density function of latent times *τ, τ*_1_ and *τ*_3_. The quantity *γ* is the total fraction of infected people that has recovered in the long term (i.e. for *t* → +∞) and *β* = 1 − *γ* is the fraction of infected people that, for *t* → +∞, has entered the deceased compartment. Also note that summing up Eq.(9-13) yields, as expected, Eq. (1). Note that in order to derive the dynamic equations of the infection diffusion we never had the need to refer to the concept of rate of removal, which was a crucial point in the Kermack and McKendrick argument (*9*). Evolution equations are, instead, naturally derived by enforcing mass conservation laws, once known the probability density functions of latent times. The present theory does not pose any restriction to the functional form of the probability density functions *p*_E_ (*τ*), *p*_EI_ (*τ, τ*_1_), *p*_EIH_ (*τ, τ*_1_, *τ*_2_), *p*_EID_ (*τ, τ*_1_, *τ*_3_). This is a great advantage as it is widely recognized that the such probability distributions very often do not obey the exponential distribution law. Attempts, indeed have been made to overcome this limitation by resorting to gamma-distributed or to the sub-class of the Erlang-distributed latent times, which allow the splitting of the problem in a sum/sequence of compartments each with a specific exponential distribution of latent times (*39–41*). However, many real latent time distributions cannot be accurately described by means of the aforementioned distributions, which that cannot either account for the statistical dependence of latent times. Recalling that *p*_E_ (*τ*) = ∫ *dτ*_1_*p*_EI_ (*τ, τ*_1_), Eqs. (3, 9, 10) show that, beside the quantity *ν* (*t*), the infection spreading is governed by the joint probability density function *p*_EI_ (*τ, τ*_1_), which needs to be accurately measured to guarantee reliable predictions and suggest effective epidemic mitigation policies. Note that the dynamics of the removed, healed and deceased compartments can be solved a posteriori, once known the distributions *p*_EIH_ (*τ, τ*_1_, *τ*_2_), *p*_EID_ (*τ, τ*_1_, *τ*_3_).

## 2 The large class of SIR/SEIR-type models as a particular case of the present theory

In this section we show that the present theory contains as a subclass the classical SIR/SEIR and similar compartmental models, which implicitly assume that latent times are exponentially distributed. So let us make the hypothesis that *p*_E_ (*τ*) = *μ* exp (−*μτ*), replacing in Eq.(9) and using integration by parts we obtain

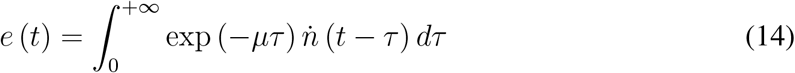

Noting that

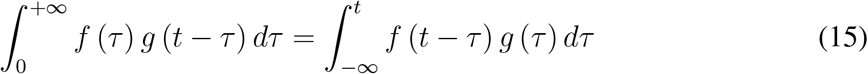

yields

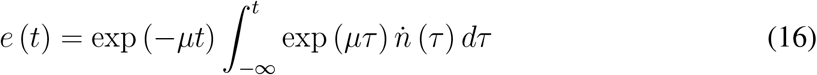

taking the time derivative and recalling Eq. (3)

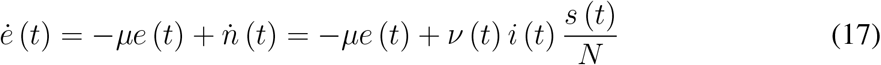

which is the classical equation for the exposed compartment in the classical SIR/SEIR model. Now let us assume that the latent times *τ* and *τ*_1_ are statistically independent i.e.

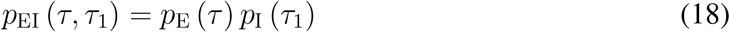

and the *τ*_1_ is exponentially distributed, then we have

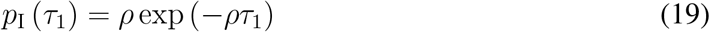

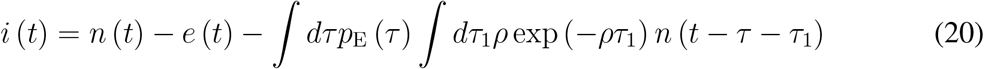

recalling Eq. (15), integration by parts yields

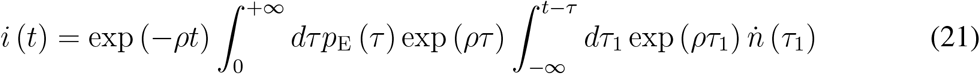

Taking the time derivative of both hand sides and recalling Eqs. (8, 17) lead to

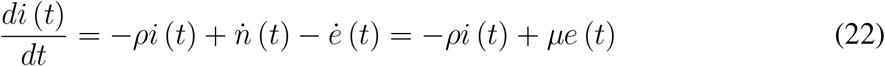

which is the classical SEIR equation for the infectious compartment. Following the same path, we now take

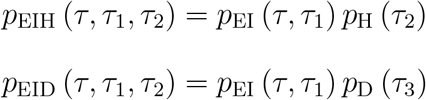

with

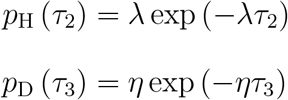

replacing in Eq. (11), using integration by parts and recalling Eq. (15) yields

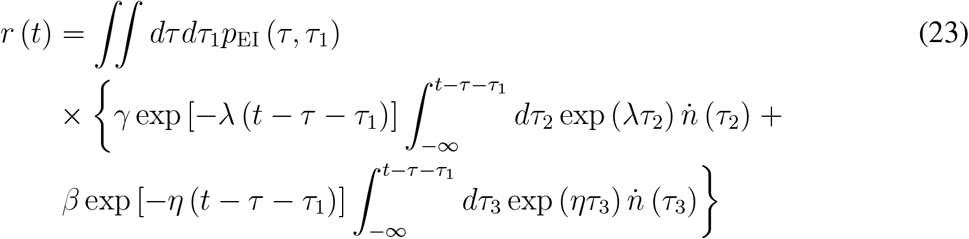

Let us introduce the auxiliary variables

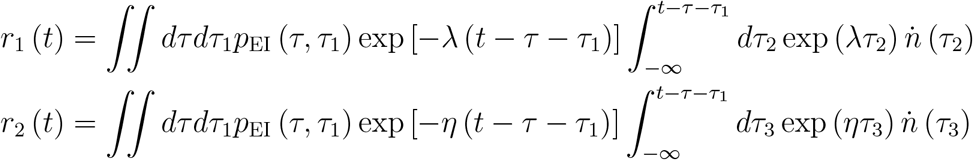

so that *r* = *γr*_1_ + *βr*_2_. Taking the time derivative and recalling Eqs. (10, 11) we get

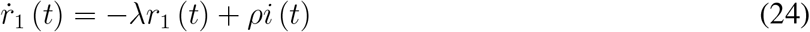

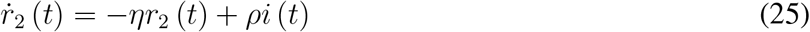

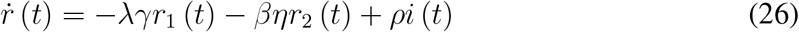

and similarly

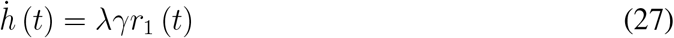

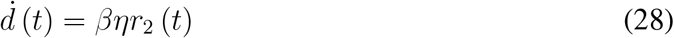

We then have shown that the entire class SIR/SEIR and similar models are a particular case of the present theory.

## 3 The basic and the effective reproduction numbers

Now let us calculate the number of secondary infections *χ* (*τ*_1_), at the beginning of infection out-break, caused by a newly infectious individual (with infectious latent time *τ*_1_) in a totally susceptible population. This number is 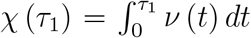, where we have assumed that the infection rate depends on time *t*. Then, the average number of secondary infections caused by a single newly infectious individual, i.e. the so called basic reproduction numbers *R*_0_, is

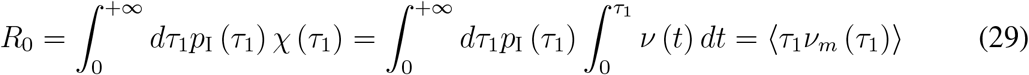

where 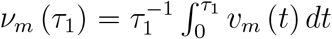 and 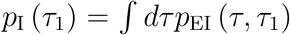 is the density probability function of latent times *τ*_1_ in the infectious compartment. The symbol ⟨ ⟩ stands for the statistical average. Note that the above definition is consistent with the usual definition in terms of survival function ℱ_I_ (*τ*_1_) = *P* (*t* ≥ *τ*_1_), where *P* (*t* ≥ *τ*_1_) is the probability for a newly infectious individual to be infectious up to time *τ*. In fact, let us observe that 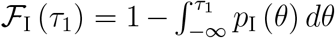, which yields

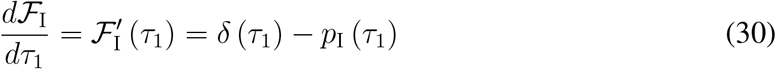

Replacing Eq. (30) in Eq. (29) we get

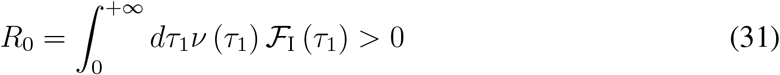

where we used integration by parts. Eq. (31) is the usual first principle formulation of the basic reproduction number (*42, 43*).

In order to monitor the evolution of infection spreading it could be useful to define an instantaneous effective reproduction number as

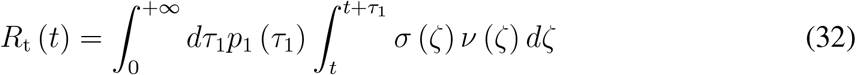

where *σ* = *s/N* = (1 − *n/N*) is the instantaneous fraction of susceptible population. Using Eq. (30) and integrating by parts one obtains

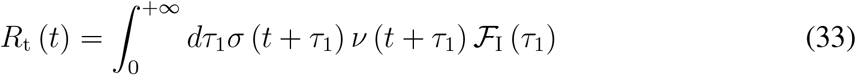

The reproduction number defined as in Eq. (33) can be also defined as the cohort reproduction number (*44*).

## 4 Deriving the Kermack and McKendrick integro-differential equations

Here we show that the present theory allows for an clear physical interpretation of the kernels of the integro-differential equations presented by Kermack and McKendrick (*9*). To this end note that Eqs. (9, 10, 11, 12, 13) can be rephrased as (see Sec. B for the detailed derivation)

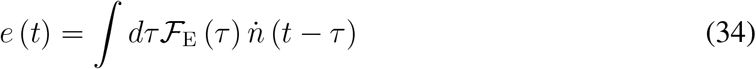

where the survival function is 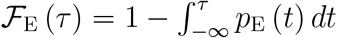 so that 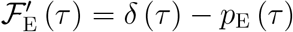, and we have used integration by parts. Following the same argument Eq. (10), can be recasted as

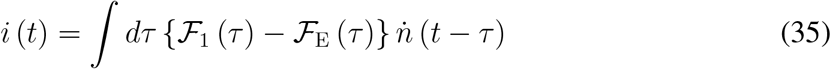

where we have changed the integration variable from *τ*_1_ to *θ*_1_ = *τ* + *τ*_1_ and used that the probability density function *p*_1_ (*θ*_1_) of the quantity *θ*_1_ is *p*_1_ (*θ*_1_) = ∫*dτp*_EI_ (*τ, θ*_1_ − *τ*). The corresponding survival function is 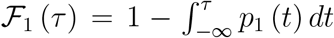. Note that *θ*_1_ is the stochastic time span from the occurrence of infection to the time of removal. Analogously we get

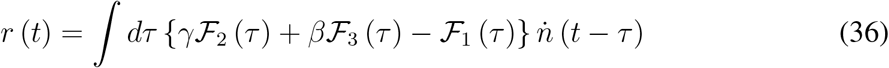

where ℱ_2_ (*θ*_2_) is the survival function of the time variable *θ*_2_ = *τ* + *τ*_1_ + *τ*_2_ and ℱ_3_ (*θ*_3_) is the survival function of *θ*_3_ = *τ* + *τ*_1_ + *τ*_3_. Now replacing Eq. (10) in Eq. (2) we get for *t >* 0

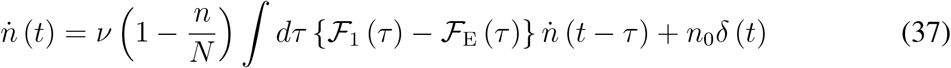

Eqs. (35, 36, 37) are formally equivalent to Eqs. (15, 14, 13) reported by Kermack and McKendrick in their paper (1927) (*9*). So we can interpret the Kernels of the Kermack and McKendrick model in terms of survival functions of latent times. Such distribution are crucial quantities which govern the infection spreading and can be measured from observed data (see for example Ref. (*45*)). Note also that for the healed and deceased compartments the following equations hold true

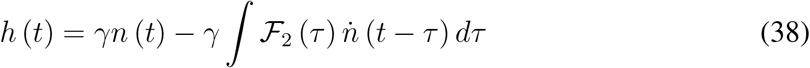

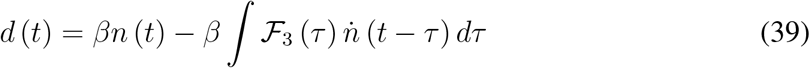

It is worth noticing that all the equations are time-invariant linear equations except for Eq. (37) which is non linear.

## 5 Linearized equations and stability analysis

In order to study the stability of the infection spreading it is enough to look at the Eq. (37) under the assumption that the ratio *n/N* ≪ 1. This allows to look at the linearized the equation to get

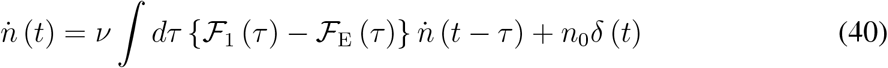

Note that ℱ_1_ (*τ*) *>* ℱ_E_ (*τ*), lim_*τ→*0_+ {ℱ_1_ (*τ*) − ℱ_E_ (*τ*)} = 0, lim_*τ→*+*∞*_ {ℱ_1_ (*τ*) − ℱ_E_ (*τ*)} = 0, and ∫*dτ* {ℱ_1_ (*τ*) − ℱ_E_ (*τ*)} = ⟨*θ*_1_⟩ − ⟨*τ* ⟩ = ⟨*τ*_1_⟩. These conditions allow us to define the probability density function *q* (*ζ*) of the positive stochastic time variable *ζ >* 0, as

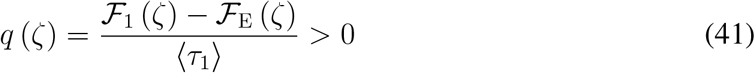

Note in fact that ∫ *q* (*ζ*) *dζ* = 1. Also observe that *q* (*ζ* = 0) = 0 and *q* (*ζ* → +∞) = 0. The moment *m*_*k*_ of *q* (*τ*) are

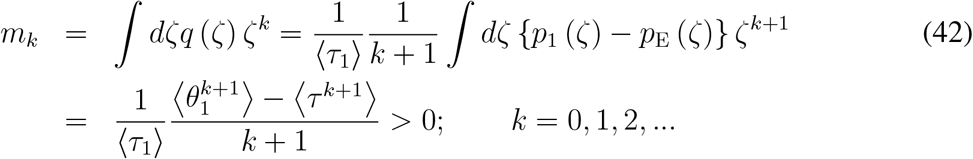

where we used integration by parts and considered that the survival function exponentially decreases at large *ζ*. The positiveness of *m*_*k*_ follows directly from the binomial theorem and considering that both *τ* and *τ*_1_ are larger than zero. Note in fact that

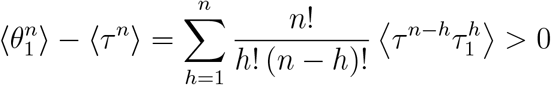

Eq. (40) can be, then, recasted in the following form

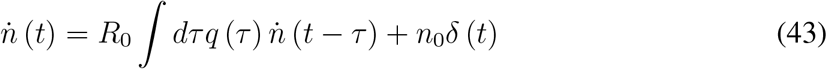

where we used that *R*_0_ = *ν* ⟨*τ*_1_⟩ or equivalently as

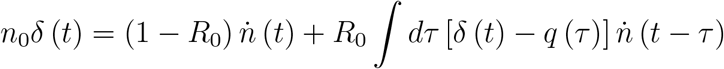

Then integrating over time *t* and recalling that 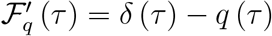,

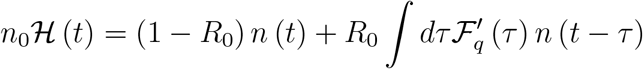

Integrating by parts and using the commutativity of the convolution product we get

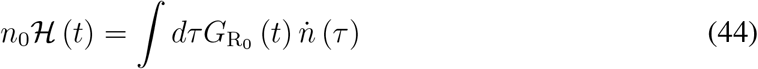

where

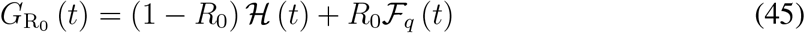

Being ℱ_q_ (*t <* 0) = 0, i.e. ℱ_q_ (*t*) satisfies the causality principle, and considering that ℱ_q_ (*τ* → +∞) = 0, ℱ_*q*_ (*τ*) can be re-interpreted, in mechanical terms, as the relaxation function of linear fluid with memory. The term (1 − *R*_0_) ℋ (*τ*) represents instead a linear spring with stiffness 1 − *R*_0_. So let us consider the linear system

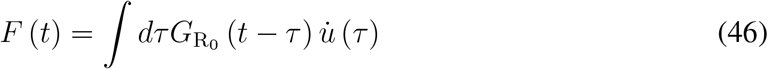

where the input is *u* (*t*) = *n* (*t*) can be interpreted as a displacement, and the output *F* (*t*) can be interpreted as a force. By taking the Laplace transform of both hand sides we have

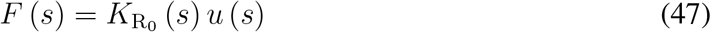

where 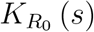 is complex stiffness of the system, i.e.

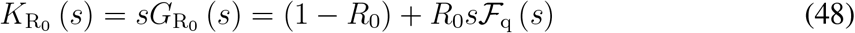

We also have

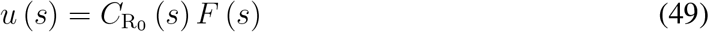

where the complex compliance is

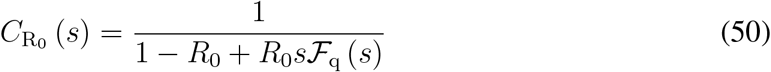

Focusing on Eq. (47) stability requires that the poles of the complex stiffness have all strictly negative real parts. This is indeed the case for the quantity *s*ℱ_q_ (*s*). In fact recalling that ℱ_q_ (*t*) satisfies causality principle, i.e., ℱ_q_ (*t <* 0) = 0, it follows that the Laplace transform of ℱ_q_ (*t*) i.e. ℱ_q_ (*s*) = ℒ [ℱ_q_ (*t*)] (*s*), is analytic in the right half of the complex plane, i.e. all the poles of ℱ_q_ (*s*) have strictly negative real parts. Regarding the spring term (1 − *R*_0_) in Eq. (48) stability also requires that the stiffness of the spring is positive, that is *R*_0_ *<* 1. One concludes that, when the controlled variable is the displacement *u* (*t*), the linear system defined by the equation 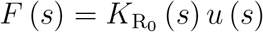 is stable provided that *R*_0_ *<* 1. In such a case it is easily shown that also the quantity

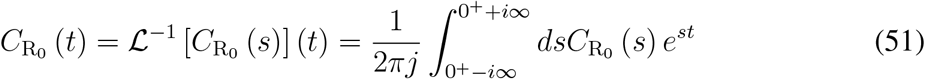

(where 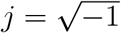) satisfies the causality principle, so that 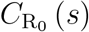 is analytic in the right half of the complex plane. Note that in terms of 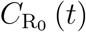 we can write

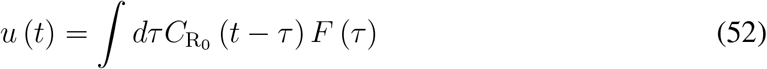

The stability of the system is then only governed by the basic reproduction number *R*_0_ and is guaranteed for 0 *< R*_0_ *<* 1. In such conditions being *F* (*t*) = *n*_0_ℋ (*t*), i.e. *F* (*s*) = *n*_0_*/s*, the epidemic will vanish in the long term with a total number of infected individuals equal to

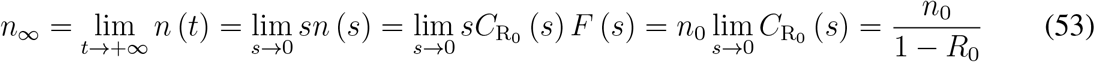

We will also have *i*_*∞*_ = *e*_*∞*_ = *r*_*∞*_ = 0 and *h*_*∞*_ = *γn*_*∞*_ and *d*_*∞*_ = *βn*_*∞*_.

## 6 The case *R*_0_ = 1

In this section we focus on what happens when *R*_0_ = *ν* ⟨*τ*_1_⟩ = 1 under the hypothesis that *n* (*t*) */N* ≪ 1. In such a case we have

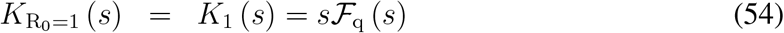

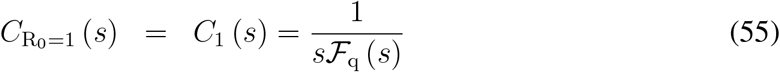

Therefore the system will behave as linear fluid with memory. Being lim_*s→*0_ *s*ℱ_q_ (*s*) = ℱ_q_ (*t* → +∞) = 0, the complex compliance *C*_1_ (*s*) has a pole at *s* = 0, all the others being in the left-half of the complex plane. Now, recalling that *s*ℱ_q_ (*s*) = 1 − *q* (*s*) and considering that

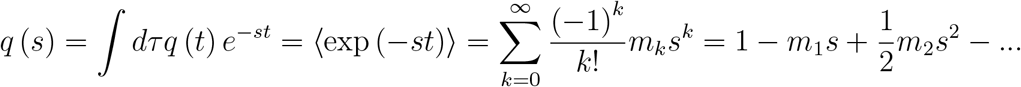

for *s* → 0 at first order in *s* we have *s*ℱ_q_ (*s*) = 1 − *q* (*s*) = *m*_1_*s*. Then, *C*_0_ (*s*) = 1*/* (*m*_1_*s*) and *u* (*s*) = *F* (*s*) */* (*m*_1_*s*). Recalling that, in our case, *F* (*t*) = *n*_0_ℋ (*t*), i.e. *F* (*s*) = *n*_0_*/s*, we get that in the long term the number of infected people *n* (*t*) will increase linearly with a rate

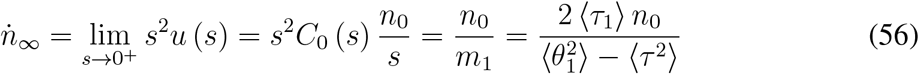

Eq. (56) shows that the first moment *m*_1_ of the probability density function *q* (*τ*) governs the velocity of spreading when *R*_0_ = 1. In the long term we also have

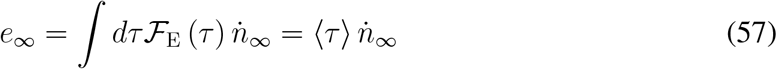

Similarly

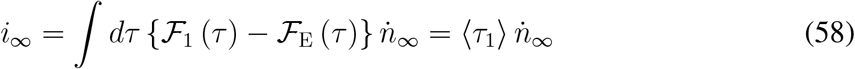

and

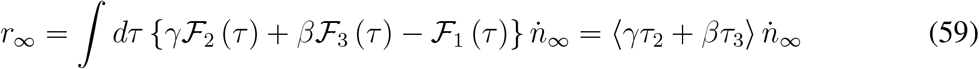

Regarding the healed individuals, taking the time derivative of

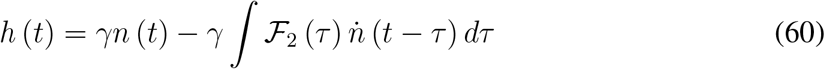

and observing that in the long term 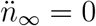 yields

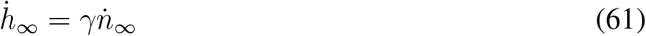

Similarly for the deceased people

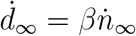

## 7 Case study: the COVID-19 epidemic in Italy

In this Section we use the proposed model to simulate the spread of COVID-19 disease in Italy during the first 120 days from the first COVID-19 outbreak in the country. As already observed, the knowledge of the infection rate *ν* (*t*) and joint probability density function *p*_EI_ (*τ, τ*_1_) is crucial to described the spreading of the disease. To estimate *p*_E_ (*τ*) we use data reported in Ref. (*45*) (for the China case), which allow an estimation of the following probability density functions: (i) *q*_IN_ (*t*_1_) where *t*_1_ is the incubation period (i.e., the time from infection to illness onset), and (ii) *q*_IR_ (*t*_2_) with *t*_2_ being the time interval from illness onset to first medical visit. In particular, assuming that on the average an individual becomes infectious *λ* days before the appearance of the symptoms, we can estimate the distribution *p*_E_ (*τ*) from *q*_IN_ (*t*_1_) by simply scaling the time axis of the quantity *a* = ⟨*t*_1_⟩^−1^ (⟨*t*_1_⟩ − *λ*), where ⟨*t*_1_⟩ = ∫*dt*_1_*t*_1_*q*_IN_ (*t*_1_), to get

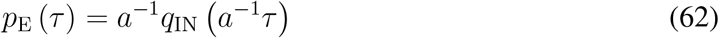

Now note that the sum of the incubation period *t*_1_ plus the period *t*_2_ from illness to first medical visit (we assume that at the first medical visit the infected individual is isolated) obeys the relation *t*_1_ + *t*_2_ = *τ* + *τ*_1_ (where *τ* is the latent time in the exposed compartment and *τ*_1_ the latent time in the infectious compartment), then we have that

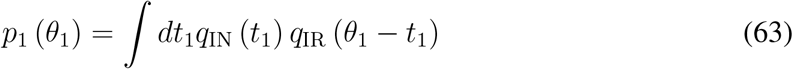

Fig. 2(a) shows the probability density function *p*_E_ (*τ*) of the latent time *τ* in the exposed compartment for *λ* = 2 days (note that researchers estimate that people who get infected with the coronavirus are most contagious 2 days before the illness onset). Fig. 2(b) reports the probability density function *p*_1_ (*θ*_1_) of the latent time *θ*_1_ = *τ* + *τ*_1_ obtained using Eq. (63).

**Figure 2:**
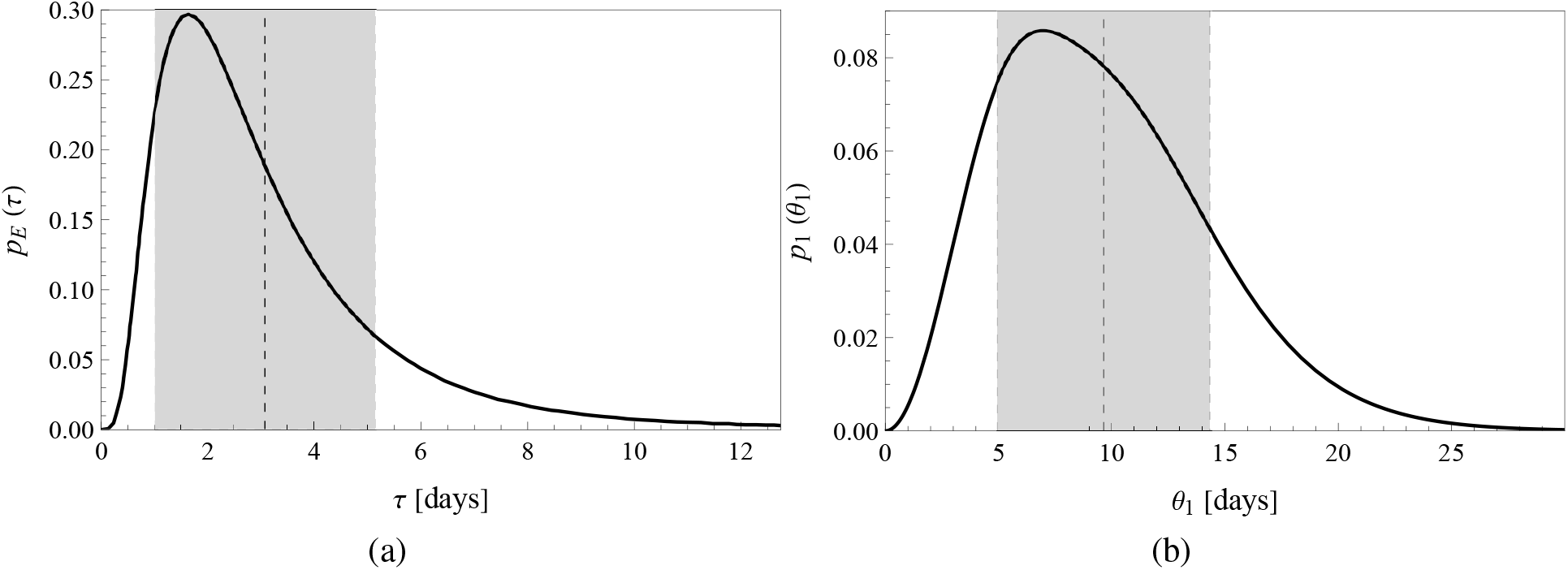
The probability density function *p*_E_ (*τ*) of the latent times *τ* (sojourn time in the exposed compartment) for *λ* = 2 days (a); the probability density function *p*_1_ (*θ*_1_) of the times *θ*_1_ = *τ* + *τ*_1_ (latency time from infection to isolation).

Now let us focus on the latent times *τ*_2_ (i.e. the sojourn time the individuals spend into the removed compartment before healing) and *τ*_3_ (i.e. the time period spent in the removed compartment before death). In this case the density probability functions *p*_H_ (*τ*_2_) and *p*_D_ (*τ*_3_) have been estimated from the data available from Refs. (*46–48*). In particular the distribution of latent time frequencies have been fitted through a double Gaussian distribution

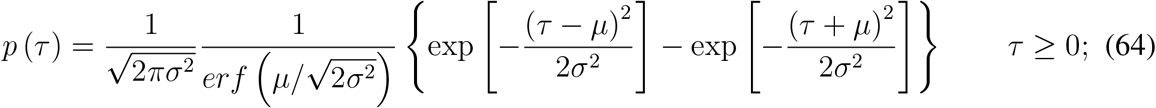

The fit parameters are *μ* and *σ*. Fig. 3(a) shows the probability density function *p*_H_ (*τ*_2_) of the latent time *τ*_2_ from isolation to recovery (*μ*_H_ = 34, *σ*_H_ = 17), whereas Fig. 2(b) reports the probability density function *p*_D_ (*τ*_3_) of the latent time *τ*_3_ from isolation to death (*μ*_D_ = 7, *σ*_D_ = 3.5). We also report the trend of the probability density functions *p*_2_ (*θ*_2_) of *θ*_2_ = *θ*_1_ + *τ*_2_ [see Fig. 4(a)] and *p*_3_ (*θ*_3_) where *θ*_3_ = *θ*_1_ + *τ*_3_ [see Fig. 4(b)]. Table 65 reports the average values and the standard deviation of the probability density function of times *τ, θ*_1_, *τ*_2_, *θ*_2_, *τ*_3_, *θ*_3_.

**Figure 3:**
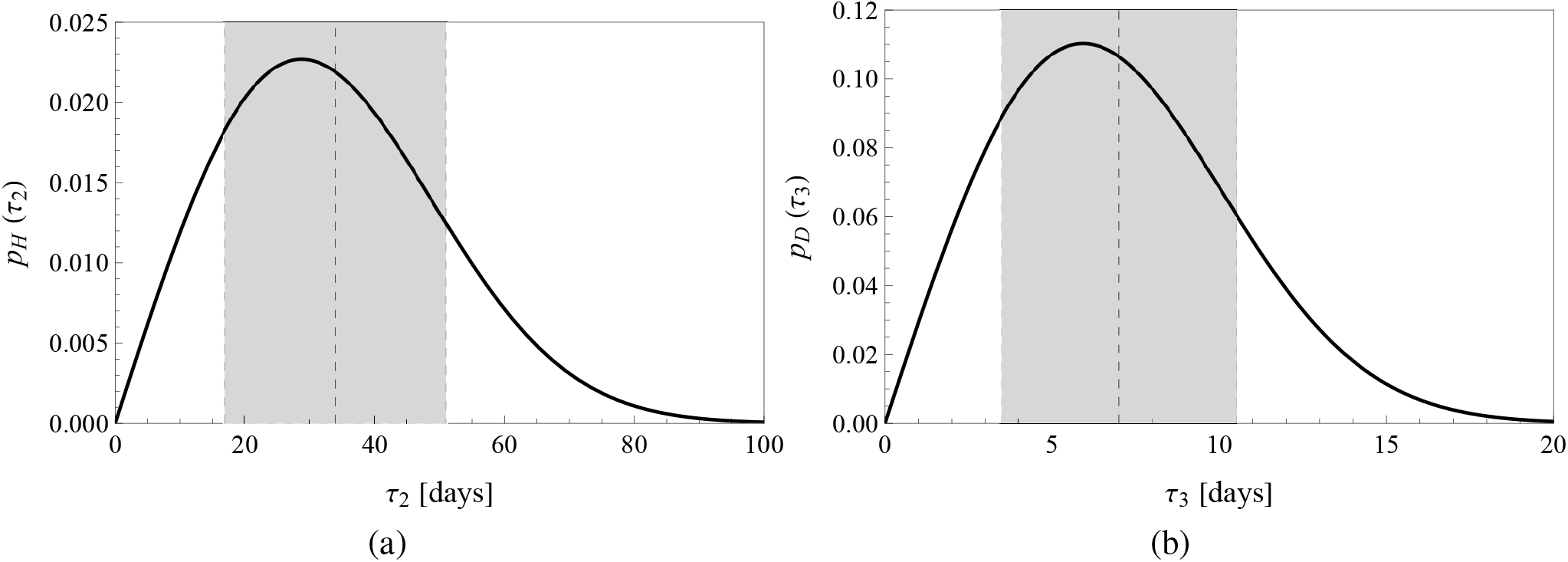
The probability density function *p*_H_ (*τ*_2_) of the latent times *τ*_2_ (latency time from isolation to healing) (a); the probability density function *p*_D_ (*τ*_3_) of the latent times *τ*_3_ (latency time from isolation to death), (b).

**Figure 4:**
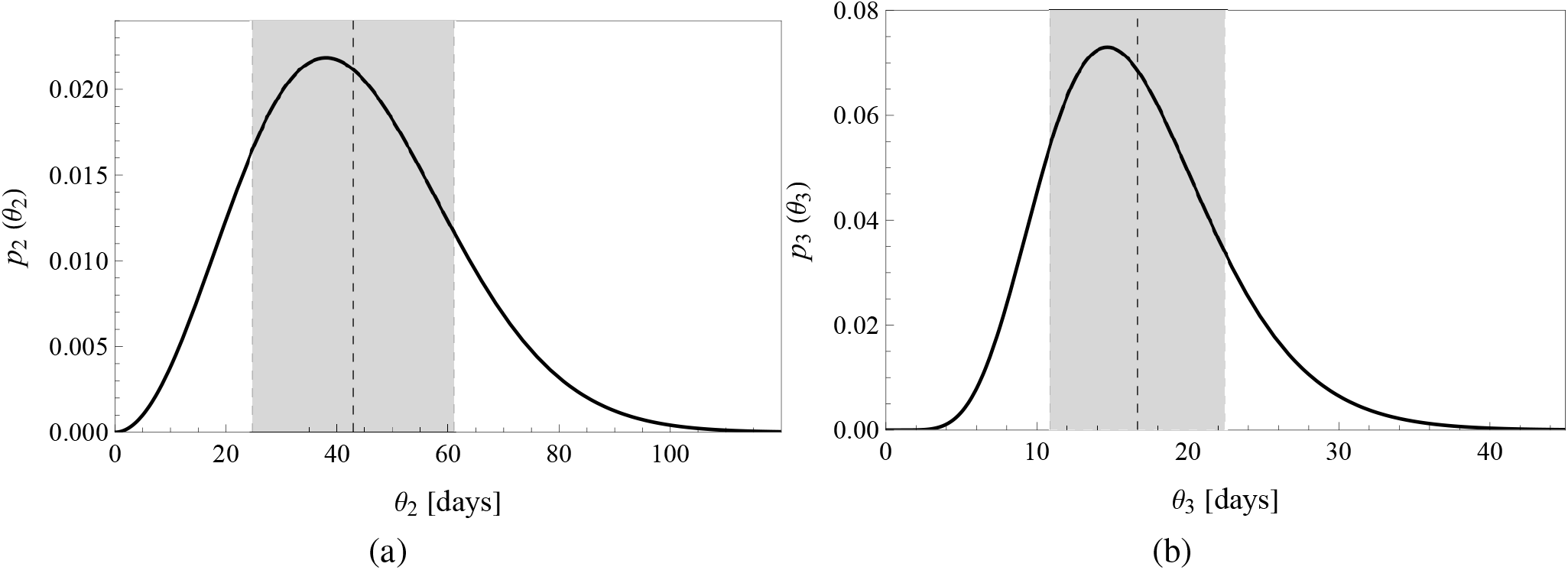
The probability density function *p*_2_ (*θ*_2_) of *θ*_2_ = *τ* + *τ*_1_ + *τ*_2_ (latency time from infection to healing) (a); the probability density function *p*_3_ (*θ*_3_) of *θ*_3_ = *τ* + *τ*_1_ + *τ*_3_ (latency time from infection to death), (b).

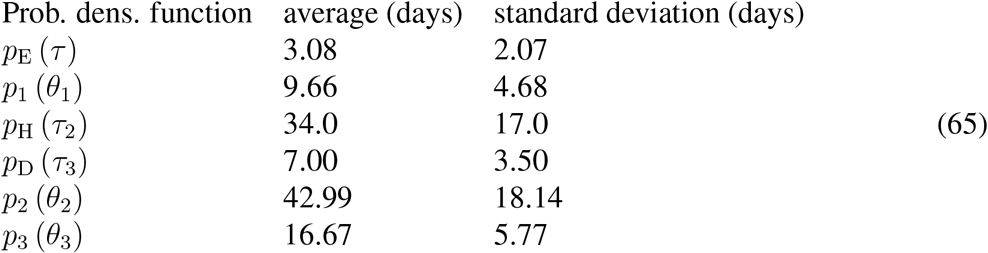

In order to simulate the effect of the control measures undertaken by the government (e.g. lockdown, quarantine and mask obligation) to limit the diffusion of the infection, we implement an exponential decrease of the infection rate *ν* = *ν* (*t*) (*49, 50*), which takes place starting from the time *T*_Q_ at which control measure are enforced, i.e.

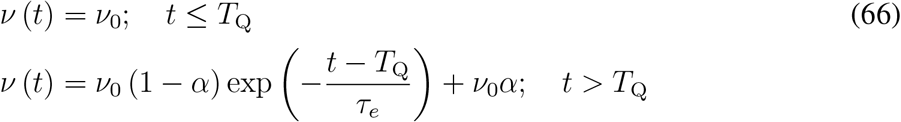

where *τ*_*e*_ is a time constant and *ν*_0_*α* is the latent infection rate originating from imperfect social distancing.

The set of parameters used to run the model for the Italy case is *n*_0_ = *n* (*t* = 0) = 117, *ν*_0_ = 0.503, *T*_Q_ = 25.7 days, *τ*_*e*_ = 10.41 days, *α* = 0.167, *γ* = 0.86, *β* = 0.14. Figure 5(a) shows the time evolution of the cumulative number *n* (*t*) of infected cases during the first 120 days from the first COVID-19 outbreak in Italy, whereas Fig. 5(b) shows the daily new infected cases 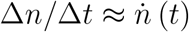 during the same period of time. Model predictions are in very good agreement with the real data, this can be accessed at the URL: https://opendatadpc.maps.arcgis.com/apps/dashboards/b0c68bce2cce478eaac82fe38d4138b1 Let us estimate the basic reproduction number *R*_0_. To this end we first consider that *p*_I_ (*τ*_1_) is vanishing small for latent times *τ*_1_ *> T*_Q_ therefore the contribution to the *τ*_1_-integral in Eq. (29) is limited to times *τ*_1_ *< T*_Q_. Thus, considering that *ν* (*t*) = *ν*_0_ for *t* ≤ *T*_Q_ we get

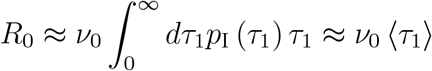

Recalling that ⟨*τ*_1_⟩ ≈ 7 days, we get *R*_0_ ≈ 3.5 which is close the value found in other studies (*51, 52*). Figure 6(a) shows the time evolution of the cumulative number *e* (*t*) of exposed cases, whereas Fig. 6(b) shows the cumulative number *i* (*t*) of individuals in the infectious compartment. Figure 7(a) shows the time evolution of the number *r* (*t*) of individuals in the removed compartment and Figure 7(b) the evolution of the daily removed individuals 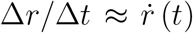. Model predictions are compared with real data. We notice a certain time-lag between the model predictions and the real data set for the removed compartment, the theoretical and real curve run, indeed, parallel to each-other. The reason for such a difference does not have a clear interpretation. However, we believe the discrepancy should be related to an asynchronous recording of the number of infected individual and the number removed ones. Figure 8(a) shows the time evolution of the number *h* (*t*) of recovered (healed) individuals and Figure 8(b) the number *d* (*t*) of the deceased individuals. Also in this case a time-lag is observed similar to the one observed for the compartment of removed individuals.

**Figure 5:**
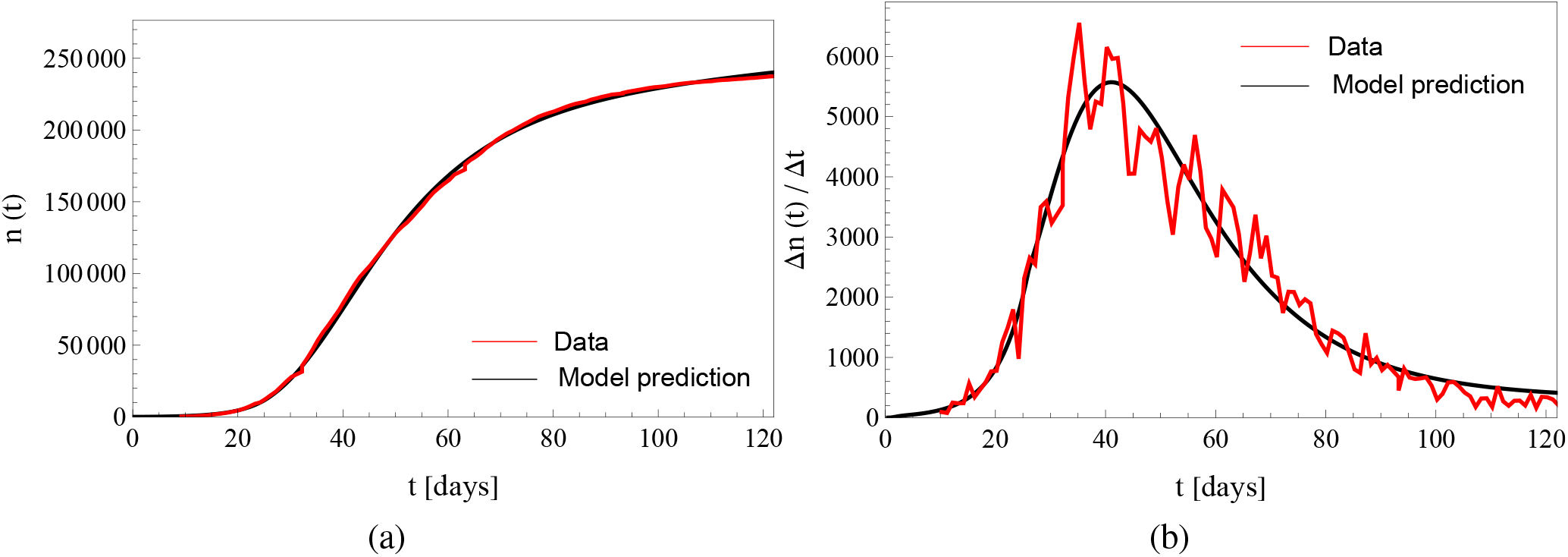
The time evolution of the cumulative number of infected cases *n* (*t*) during the first 120 days of the first COVID outbreak in Italy, (a); the daily new infected cases 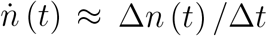 during the same period of time, (b).

**Figure 6:**
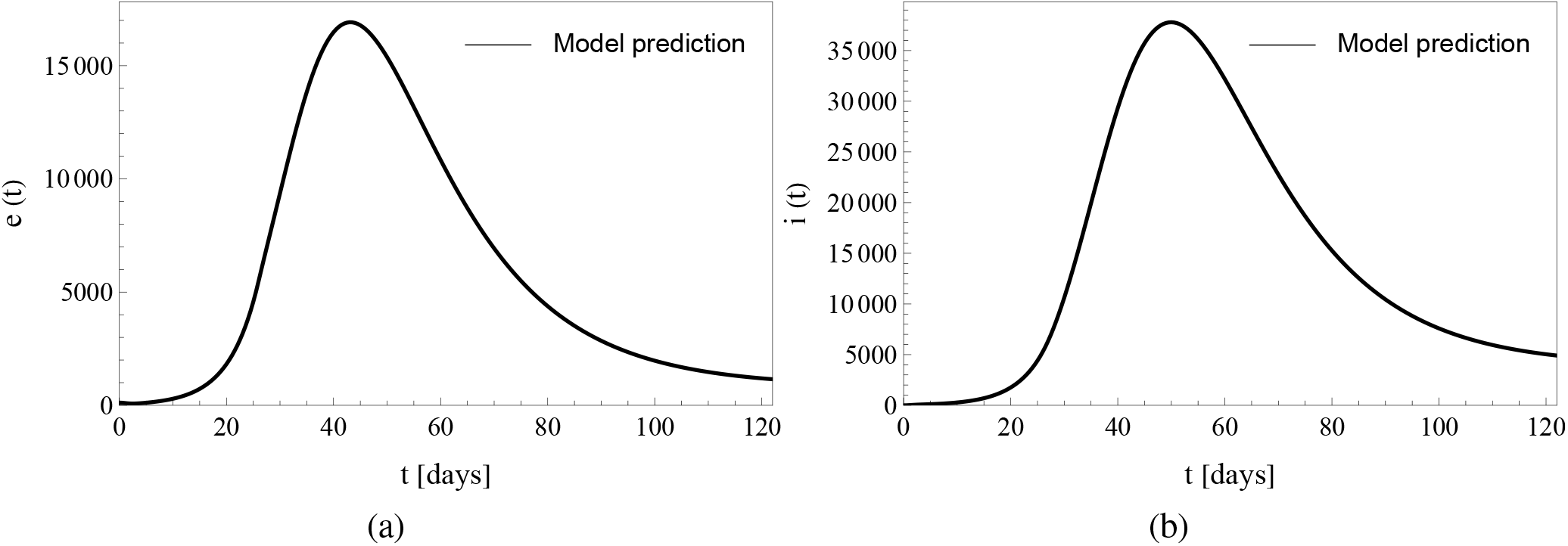
The time evolution of the number of individuals in the exposed compartment *e* (*t*), (a); The time evolution of the number of individuals in the infectious compartment *i* (*t*), (b).

**Figure 7:**
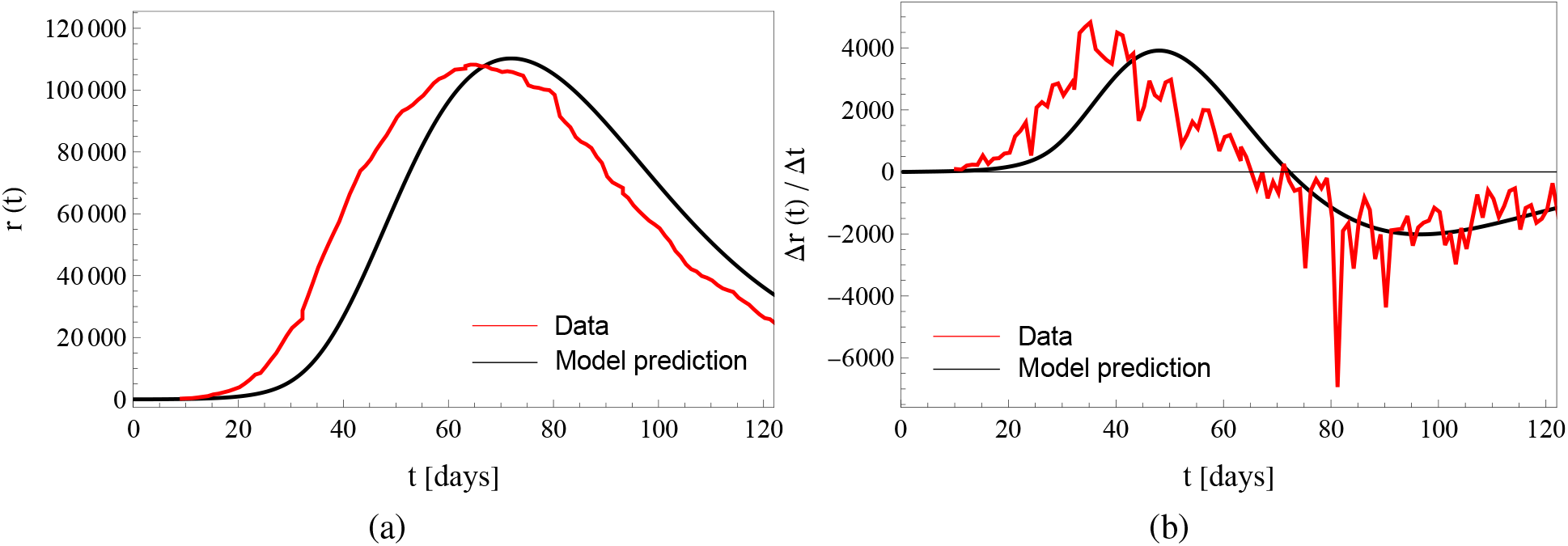
The time evolution of individuals in the removed compartment *r* (*t*), (a); the time evolution of the daily removed individuals 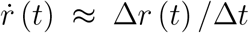, (b). Model predictions are compared with real data.

**Figure 8:**
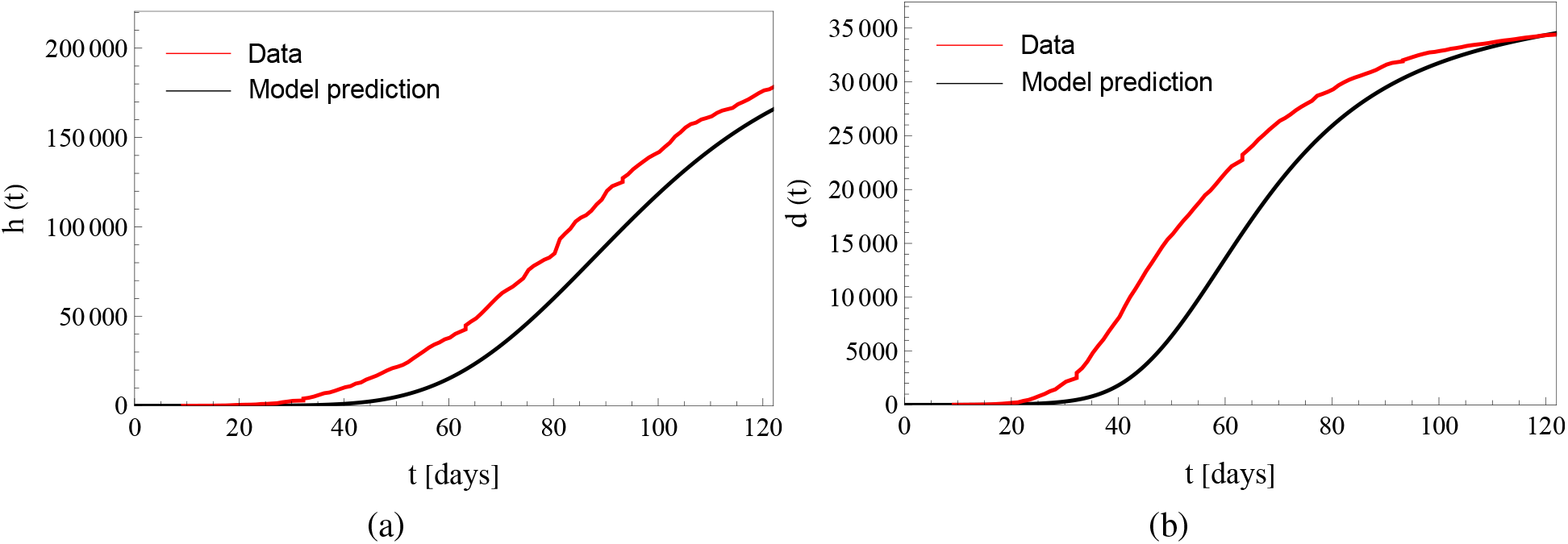
The time evolution of healed individuals *h* (*t*), (a); the time evolution of deaths *d* (*t*), (b). Model predictions are compared with real data.

## 8 Conclusion

We present a general theory of infection dynamics, leading to Volterra like equations where the kernels have a clear physical interpretation in terms of probability density function or in terms of survival function of latent time distributions, thus overcoming the main difficulty of Kermack and McKendrick model (1927), where the interpretation of the physical meaning of the kernels was really unclear and difficult to interpret in terms of real data. Our theory contains as a particular case the whole class of SIR, SEIR. Beside the infection rate *ν* (*t*), a very central role is played by the joint probability density function *p*_EI_ (*τ, τ*_1_) of the latent times *τ* and *τ*_1_ the individuals spend in the exposed and infectious compartments respectively. The quantity *p*_EI_ (*τ, τ*_1_) is then a crucial information needed to predict how the infection will develop in time. The theory does not pose any restriction to the functional form of the probability density functions of compartmental latent times, which usually do not obey the exponential distribution, as, instead, implicitly assumed in classical SIR and SEIR-type models. When the number of infections is negligible with the entire population, the equations can be solved in general and their stability studied by resorting their representation in the Laplace domain. The particular case of *R*_0_ = 1 is also investigated to show that the moments of the probability density functions of latent times, govern the asymptotic infection rate in this case. The present theory has been employed to simulate the spreading of COVID-19 infection in Italy during the first 120 days subsequent to the identification of the first cases. We found a very good agreement between theoretical predictions and real data. Our estimation of the basic reproduction number is *R*_0_ ≈ 3.5 in agreement with other studies.

### A Compartmental dynamic equations

Consider the number *δm*_*k*_ of exposed individuals with latent time *τ*_*k*_ which enters, during a time interval *δt*, the infectious compartment at time *t*. Of such newly infectious individuals a certain number *δl*_*kh*_, will leave the infectious department after a latent time 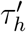, the fraction *δl*_*kh*_*/δm*_*k*_ is

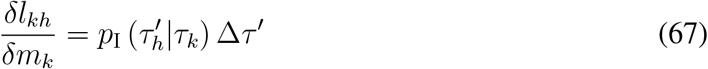

where *p*_I_ (*τ* ^*′*^|*τ*) is the conditional probability density function of the infectious latent times *τ* ^*′*^ given the exposed latent time *τ*. Therefore the increase *δi*_*k*_ of infectious individuals at time *t* due to the contribution of exposed individuals *δm*_*k*_ with latent time *τ*_*k*_ is

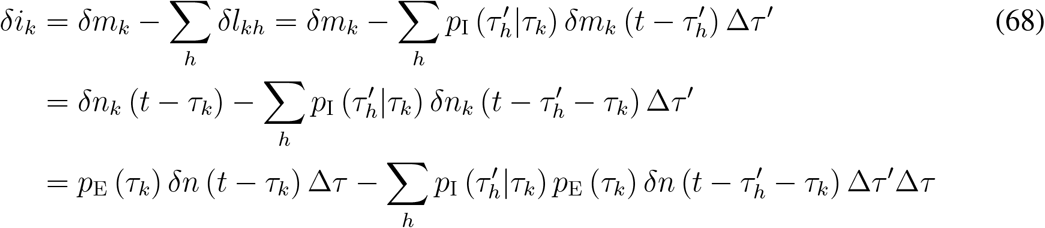

and summing up over the index *k*

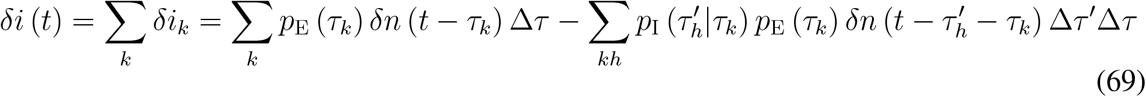

or

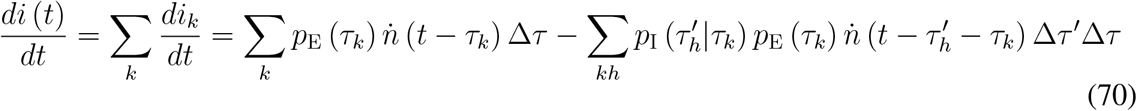

Taking the limit of vanishing Δ*τ* and Δ*τ* ^*′*^ gives

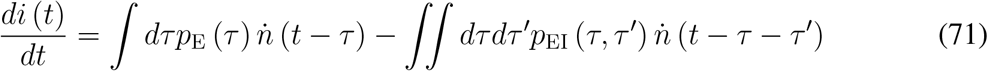

where we have used that *p*_I_ (*τ* ^*′*^|*τ*) *p*_E_ (*τ*) = *p*_EI_ (*τ, τ* ^*′*^), with *τ* and *τ* ^*′*^ both greater or equal to zero. Integrating over time *t* and renaming *τ* ^*′*^ → *τ*_1_ give

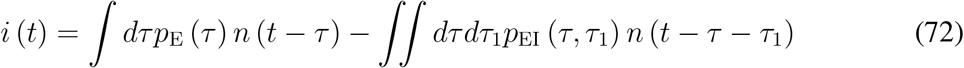

Recalling Eq. (9) we then get Eq. (3).

A similar argument applies to the removed compartment. Let us consider *δl*_*kh*_ individuals with latent time *τ*_*k*_ and 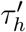 entering, during the time step *δt*, the removed compartment at time *t*. Of such individuals a number 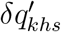 will recover after a latent time 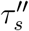 and another number 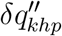 with latent time 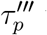 will enter the deceased compartment. We now consider that in the long term a fraction *γ* of the individuals will recover and the fraction *β* = 1 − *γ* will move into the deceased compartment. The we can write

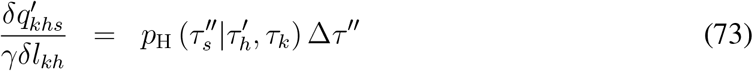

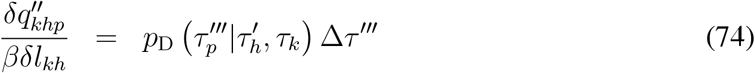

therefore the increase *δr*_*kh*_ of individuals in the removed compartment at time *t* due to the contribution of infectious individuals *δl*_*kh*_ with latent times *τ*_*k*_ and 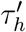 is

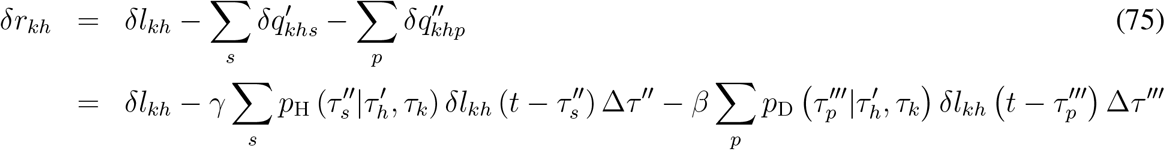

consider now that 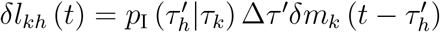 and 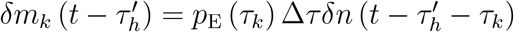. Then, recalling that 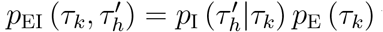 we have 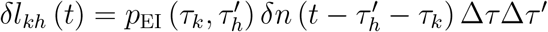, and replacing into the above equation we get

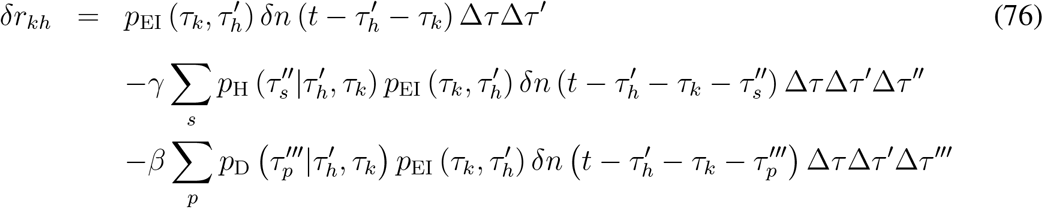

Also recalling that 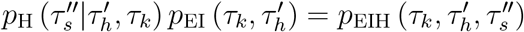 and 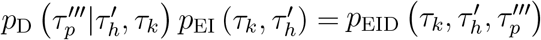 we obtain

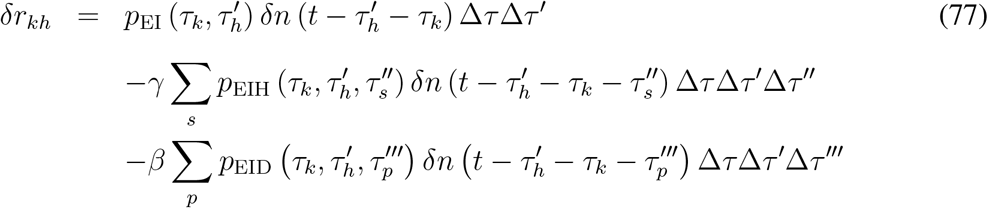

then by taking the sum over the indexes *k* and *h* and in the limit of vanishing Δ*τ*, Δ*τ* ^*′*^, Δ*τ* ^*′′*^, Δ*τ* ^*′′′*^ one obtains

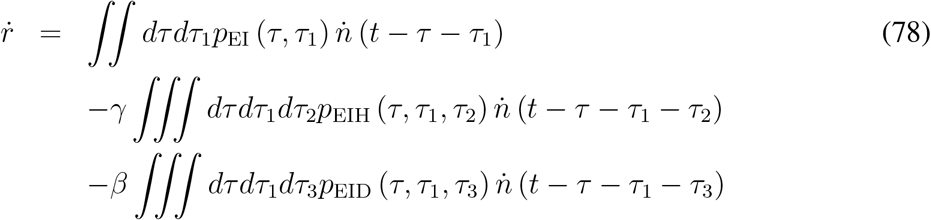

### B Rephrasing the equations

Consider Eq. (9), which describes the evolution of the number of individuals in the exposed compartment. It can be rephrased as

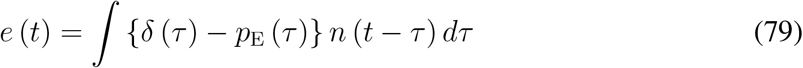

and recalling that 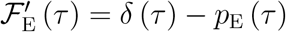 we also have

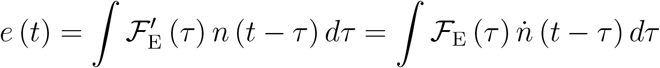

where we use integration by parts and considered that the survival function ℱ_E_ (*τ*) is zero for *τ <* 0 and decreases exponentially as *τ* → +∞. Now consider Eq (10)

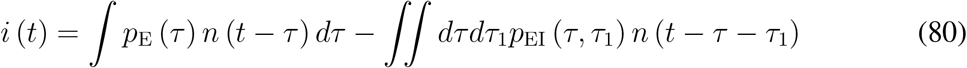

By intoducing the new integration variable *θ*_1_ such that *θ*_1_ = *τ* + *τ*_1_, the double integral at the right hand side of the equation can be rephrased as ∫ ∫*dτdθ*_1_*p*_EI_ (*τ, θ*_1_ − *τ*) *n* (*t* − *θ*_1_) and recalling that ∫*dτp*_EI_ (*τ, θ*_1_ − *τ*) = *p*_1_ (*θ*_1_) one gets

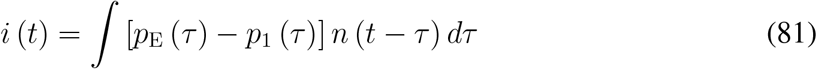

Moving to the survival functions ℱ_1_ (*τ*) and ℱ_E_ (*τ*) of the probability density functions *p*_E_ (*τ*) and *p*_1_ (*τ*) respectively we get

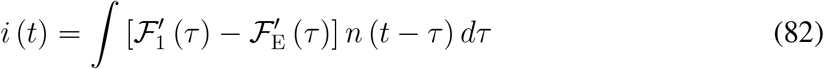

and integrating by parts

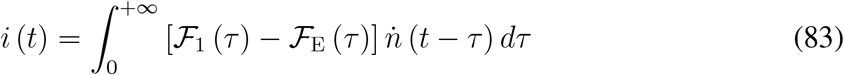

Then Eq. (3) can be rephrases as

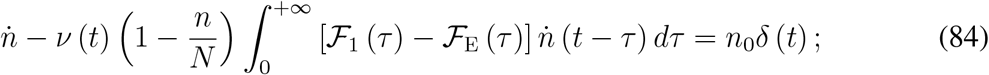

Using the same argument and setting *θ*_2_ = *τ* + *τ*_1_ + *τ*_2_ we also have

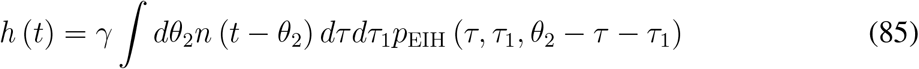

and recalling that

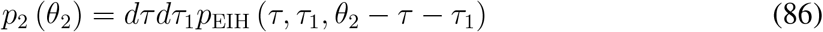

we get

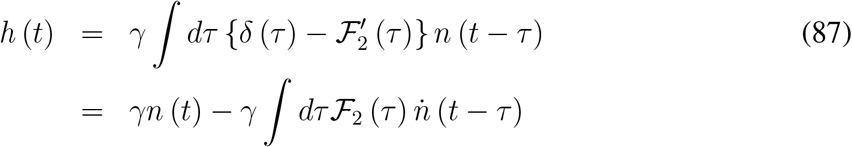

where ℱ_2_ (*θ*_2_) is the survival function of *p*_2_ (*θ*_2_). Analogously we have

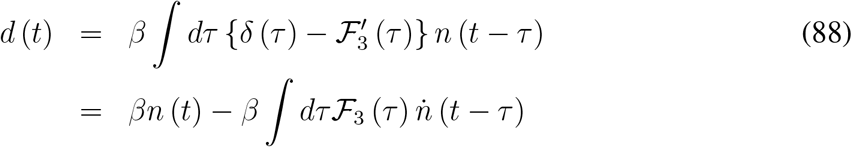

where ℱ_3_ (*θ*_3_) is the survival function of *p*_3_ (*θ*_3_), where *θ*_3_ = *τ* + *τ*_1_ + *τ*_3_.

## Supporting information

Real Data Covid Italy

Real Data Covid Italy

## Data Availability

All data produced in the present study are available upon reasonable request to the authors

