## Supplementary material for "A general theory for infectious disease dynamics": Real Data Covid Italy

url: https://opendatadpc.maps.arcgis.com/apps/dashboards/b0c68bce2cce478eaac82fe38d4138b1

| da a | stato | ricoverati_con_sintomi | terapia_intensiva | totale_ospedalizzati | isolamento_domiciliare | totale_positivi | variazione_totale_positivi | nuovi_positivi | dimessi_guariti | deceduti | casi_da_sosp | casi_da_screening | totale casi ricalc | totale_casi | tamponi | casi_testat | note |
|---|---|---|---|---|---|---|---|---|---|---|---|---|---|---|---|---|---|
| 24/02/2020 18:00 | ITA | 101 | 26 | 127 | 94 | 221 | 0 | 221 | 1 | 7 | | | 229 | 229 | 4324 | | |
| 25/02/2020 18:00 | ITA | 114 | 35 | 150 | 162 | 311 | 90 | 93 | 1 | 10 | | | 322 | 322 | 8623 | | |
| 26/02/2020 18:00 | ITA | 128 | 36 | 164 | 221 | 385 | 74 | 78 | 3 | 12 | | | 400 | 400 | 9587 | | |
| 27/02/2020 18:00 | ITA | 248 | 56 | 304 | 284 | 588 | 203 | 250 | 45 | 17 | | | 650 | 650 | 12014 | | |
| 28/02/2020 18:00 | ITA | 345 | 64 | 409 | 412 | 821 | 233 | 238 | 46 | 21 | | | 888 | 888 | 15695 | | |
| 29/02/2020 18:00 | ITA | 401 | 105 | 506 | 543 | 1049 | 228 | 240 | 50 | 29 | | | 1128 | 1128 | 18661 | | |
| 01/03/2020 18:00 | ITA | 639 | 140 | 779 | 798 | 1577 | 528 | 566 | 83 | 34 | | | 1694 | 1694 | 21127 | | |
| 02/03/2020 18:00 | ITA | 742 | 166 | 908 | 927 | 1835 | 258 | 342 | 149 | 52 | | | 2036 | 2036 | 23345 | | |
| 03/03/2020 18:00 | ITA | 1034 | 229 | 1263 | 1000 | 2263 | 428 | 466 | 160 | 79 | | | 2502 | 2502 | 25856 | | |
| 04/03/2020 18:00 | ITA | 1346 | 295 | 1641 | 1065 | 2706 | 443 | 587 | 276 | 107 | | | 3089 | 3089 | 29837 | | |
| 05/03/2020 18:00 | ITA | 1790 | 351 | 2141 | 1155 | 3296 | 590 | 769 | 414 | 148 | | | 3858 | 3858 | 32362 | | |
| 06/03/2020 18:00 | ITA | 2394 | 462 | 2856 | 1060 | 3916 | 620 | 778 | 523 | 197 | | | 4636 | 4636 | 36359 | | |
| 07/03/2020 18:00 | ITA | 2651 | 567 | 3218 | 1843 | 5061 | 1145 | 1247 | 589 | 233 | | | 5883 | 5883 | 42062 | | pd-IT-0003 |
| 08/03/2020 18:00 | ITA | 3557 | 650 | 4207 | 2180 | 6387 | 1326 | 1492 | 622 | 366 | | | 7375 | 7375 | 49937 | | |
| 09/03/2020 18:00 | ITA | 4316 | 733 | 5049 | 2936 | 7985 | 1598 | 1797 | 724 | 463 | | | 9172 | 9172 | 53826 | | |
| 10/03/2020 18:00 | ITA | 5038 | 877 | 5915 | 2599 | 8514 | 529 | 977 | 1004 | 631 | | | 10149 | 10149 | 60761 | | pd-IT-0005 |
| 11/03/2020 18:00 | ITA | 5838 | 1028 | 6866 | 3724 | 10590 | 2076 | 2313 | 1045 | 827 | | | 12462 | 12462 | 73154 | | nd-IT-0002 |
| 12/03/2020 18:00 | ITA | 6650 | 1153 | 7803 | 5036 | 12839 | 2249 | 2651 | 1258 | 1016 | | | 15113 | 15113 | 86011 | | |
| 13/03/2020 18:00 | ITA | 7426 | 1328 | 8754 | 6201 | 14955 | 2116 | 2547 | 1439 | 1266 | | | 17660 | 17660 | 97488 | | |
| 14/03/2020 18:00 | ITA | 8372 | 1518 | 9890 | 7860 | 17750 | 2795 | 3497 | 1966 | 1441 | | | 21157 | 21157 | 109170 | | |
| 15/03/2020 18:00 | ITA | 9663 | 1672 | 11335 | 9268 | 20603 | 2853 | 3590 | 2335 | 1809 | | | 24747 | 24747 | 124899 | | |
| 16/03/2020 18:00 | ITA | 11025 | 1851 | 12876 | 10197 | 23073 | 2470 | 3233 | 2749 | 2158 | | | 27980 | 27980 | 137962 | | nd-IT-0004;nd-IT-0006 |
| 18/03/2020 18:00 | ITA | 12894 | 2060 | 14954 | 11108 | 26062 | 2989 | 3526 | 2941 | 2503 | | | 31506 | 31506 | 148657 | | nd-IT-0009 |
| 18/03/2020 18:00 | ITA | 14363 | 2257 | 16620 | 12090 | 28710 | 2648 | 4207 | 4025 | 2978 | | | 35713 | 35713 | 165541 | | nd-IT-0012;nd-IT-0014 |
| 19/03/2020 18:00 | ITA | 15757 | 2498 | 18255 | 14935 | 33190 | 4480 | 5322 | 4440 | 3405 | | | 41035 | 41035 | 182777 | | |
| 20/03/2020 18:00 | ITA | 16020 | 2655 | 18675 | 19185 | 37860 | 4670 | 5986 | 5129 | 4032 | | | 47021 | 47021 | 206886 | | |
| 21/03/2020 18:00 | ITA | 17708 | 2857 | 20565 | 22116 | 42681 | 4821 | 6557 | 6072 | 4825 | | | 53578 | 53578 | 233222 | | |
| 22/03/2020 18:00 | ITA | 19846 | 3009 | 22855 | 23783 | 46638 | 3957 | 5560 | 7024 | 5476 | | | 59138 | 59138 | 258402 | | |
| 23/03/2020 18:00 | ITA | 20692 | 3204 | 23896 | 26522 | 50418 | 3780 | 4789 | 7432 | 6077 | | | 63927 | 63927 | 275468 | | |
| 24/03/2020 18:00 | ITA | 21937 | 3396 | 25333 | 28697 | 54030 | 3612 | 5249 | 8326 | 6820 | | | 69176 | 69176 | 296964 | | |
| 25/03/2020 18:00 | ITA | 23112 | 3489 | 26601 | 30920 | 57521 | 3491 | 5210 | 9362 | 7503 | | | 74386 | 74386 | 324445 | | |
| 26/03/2020 18:00 | ITA | 24753 | 3612 | 28365 | 33648 | 62013 | 4492 | 6153 | 10361 | 8165 | | | 80539 | 80539 | 361060 | | pd-IT-0007 |
| 27/03/2020 18:00 | ITA | 26029 | 3732 | 29761 | 36653 | 66414 | 4401 | 5959 | 10950 | 9134 | | | 86498 | 86498 | 394079 | | |
| 28/03/2020 18:00 | ITA | 26676 | 3856 | 30532 | 39533 | 70065 | 3651 | 5974 | 12384 | 10023 | | | 92472 | 92472 | 429526 | | |
| 29/03/2020 18:00 | ITA | 27386 | 3906 | 31292 | 42588 | 73880 | 3815 | 5217 | 13030 | 10779 | | | 97689 | 97689 | 454030 | | pd-IT-0009 |
| 30/03/2020 18:00 | ITA | 27795 | 3981 | 31776 | 43752 | 75528 | 1648 | 4050 | 14620 | 11591 | | | 101739 | 101739 | 477359 | | |
| 31/03/2020 18:00 | ITA | 28192 | 4023 | 32215 | 45420 | 77635 | 2107 | 4053 | 15729 | 12428 | | | 105792 | 105792 | 506968 | | |
| 01/04/2020 18:00 | ITA | 28403 | 4035 | 32438 | 48134 | 80572 | 2937 | 4782 | 16847 | 13155 | | | 110574 | 110574 | 541423 | | |
| 02/04/2020 18:00 | ITA | 28540 | 4053 | 32593 | 50456 | 83049 | 2477 | 4668 | 18278 | 13915 | | | 115242 | 115242 | 581232 | | |
| 03/04/2020 18:00 | ITA | 28741 | 4068 | 32809 | 52579 | 85388 | 2339 | 4585 | 19758 | 14681 | | | 119827 | 119827 | 619849 | | |
| 04/04/2020 18:00 | ITA | 29010 | 3994 | 33004 | 55270 | 88274 | 2886 | 4805 | 20996 | 15362 | | | 124632 | 124632 | 657224 | | |
| 05/04/2020 18:00 | ITA | 28949 | 3977 | 32926 | 58320 | 91246 | 2972 | 4316 | 21815 | 15887 | | | 128948 | 128948 | 691461 | | |
| 06/04/2020 18:00 | ITA | 28976 | 3898 | 32874 | 60313 | 93187 | 1941 | 3599 | 22837 | 16523 | | | 132547 | 132547 | 721732 | | |
| 07/04/2020 18:00 | ITA | 28718 | 3792 | 32510 | 61557 | 94067 | 880 | 3039 | 24392 | 17127 | | | 135586 | 135586 | 755445 | | |
| 08/04/2020 18:00 | ITA | 28485 | 3693 | 32178 | 63084 | 95262 | 1195 | 3836 | 26491 | 17669 | | | 139422 | 139422 | 807125 | | |
| 09/04/2020 18:00 | ITA | 28399 | 3605 | 32004 | 64873 | 96877 | 1615 | 4204 | 28470 | 18279 | | | 143626 | 143626 | 853369 | | |
| 10/04/2020 18:00 | ITA | 28242 | 3497 | 31739 | 66534 | 98273 | 1396 | 3951 | 30455 | 18849 | | | 147577 | 147577 | 906864 | | pd-IT-0011 |
| 11/04/2020 18:00 | ITA | 28144 | 3381 | 31525 | 68744 | 100269 | 1996 | 4694 | 32534 | 19468 | | | 152271 | 152271 | 963473 | | |
| 12/04/2020 18:00 | ITA | 27847 | 3343 | 31190 | 71063 | 102253 | 1984 | 4092 | 34211 | 19899 | | | 156363 | 156363 | 1010193 | | dc-IT-0003 |
| 13/04/2020 18:00 | ITA | 28023 | 3260 | 31283 | 72333 | 103616 | 1363 | 3153 | 35435 | 20465 | | | 159516 | 159516 | 1046910 | | |
| 14/04/2020 18:00 | ITA | 28011 | 3186 | 31197 | 73094 | 104291 | 675 | 2972 | 37130 | 21067 | | | 162488 | 162488 | 1073689 | | |
| 15/04/2020 18:00 | ITA | 27643 | 3079 | 30722 | 74696 | 105418 | 1127 | 2667 | 38092 | 21645 | | | 165155 | 165155 | 1117404 | | dc-IT-0005 |
| 16/04/2020 18:00 | ITA | 26893 | 2936 | 29829 | 76778 | 106607 | 1189 | 3786 | 40164 | 22170 | | | 168941 | 168941 | 1178403 | | |
| 18/04/2020 18:00 | ITA | 25786 | 2812 | 28598 | 78364 | 106962 | 355 | 3493 | 42727 | 22745 | | | 172434 | 172434 | 1244108 | | |
| 18/04/2020 18:00 | ITA | 25007 | 2733 | 27740 | 80031 | 107771 | 809 | 3491 | 44927 | 23227 | | | 175925 | 175925 | 1305833 | | |
| 19/04/2020 18:00 | ITA | 25033 | 2635 | 27668 | 80589 | 108257 | 486 | 3047 | 47055 | 23660 | | | 178972 | 178972 | 1356541 | 935310 | |
| 20/04/2020 18:00 | ITA | 24906 | 2573 | 27479 | 80758 | 108237 | -20 | 2256 | 48877 | 24114 | | | 181228 | 181228 | 1398024 | 943151 | dc-IT-0007 |
| 21/04/2020 18:00 | ITA | 24134 | 2471 | 26605 | 81104 | 107709 | -528 | 2729 | 51600 | 24648 | | | 183957 | 183957 | 1450150 | 971246 | pd-IT-0013 |
| 22/04/2020 18:00 | ITA | 23805 | 2384 | 26189 | 81510 | 107699 | -10 | 3370 | 54543 | 25085 | | | 187327 | 187327 | 1513251 | 1015494 | |
| 23/04/2020 18:00 | ITA | 22871 | 2267 | 25138 | 81710 | 106848 | -851 | 2646 | 57576 | 25549 | | | 189973 | 189973 | 1579909 | 1052577 | pd-IT-0015;pd-IT-0017 |
| 24/04/2020 18:00 | ITA | 22068 | 2173 | 24241 | 82286 | 106527 | -321 | 3021 | 60498 | 25969 | | | 192994 | 192994 | 1642356 | 1147850 | dc-IT-0009;dc-IT-0011 |
| 25/04/2020 18:00 | ITA | 21533 | 2102 | 23635 | 82212 | 105847 | -680 | 2357 | 63120 | 26384 | | | 195351 | 195351 | 1707743 | 1186526 | |
| 26/04/2020 18:00 | ITA | 21372 | 2009 | 23381 | 82722 | 106103 | 256 | 2324 | 64928 | 26644 | | | 197675 | 197675 | 1757659 | 1210639 | dc-IT-0013 |
| 27/04/2020 18:00 | ITA | 20353 | 1956 | 22309 | 83504 | 105813 | -290 | 1739 | 66624 | 26977 | | | 199414 | 199414 | 1789662 | 1237317 | |
| 28/04/2020 18:00 | ITA | 19723 | 1863 | 21586 | 83619 | 105205 | -608 | 2091 | 68941 | 27359 | | | 201505 | 201505 | 1846934 | 1274871 | |
| 29/04/2020 18:00 | ITA | 19210 | 1795 | 21005 | 83652 | 104657 | -548 | 2086 | 71252 | 27682 | | | 203591 | 203591 | 1910761 | 1313460 | |
| 30/04/2020 18:00 | ITA | 18149 | 1694 | 19843 | 81708 | 101551 | -3106 | 1872 | 75945 | 27967 | | | 205463 | 205463 | 1979217 | 1354901 | |
| 01/05/2020 18:00 | ITA | 17569 | 1578 | 19147 | 81796 | 100943 | -608 | 1965 | 78249 | 28236 | | | 207428 | 207428 | 2053425 | 1398633 | dc-IT-0015 |
| 02/05/2020 18:00 | ITA | 17357 | 1539 | 18896 | 81808 | 100704 | -239 | 1900 | 79914 | 28710 | | | 209328 | 209328 | 2108837 | 1429864 | dc-IT-0017 |
| 03/05/2020 18:00 | ITA | 17242 | 1501 | 18743 | 81436 | 100179 | -525 | 1389 | 81654 | 28884 | | | 210717 | 210717 | 2153772 | 1456911 | |
| 04/05/2020 18:00 | ITA | 16823 | 1479 | 18302 | 81678 | 99980 | -199 | 1221 | 82879 | 29079 | | | 211938 | 211938 | 2191403 | 1479910 | dc-IT-0019 |
| 05/05/2020 18:00 | ITA | 16270 | 1427 | 17697 | 80770 | 98467 | -1513 | 1075 | 85231 | 29315 | | | 213013 | 213013 | 2246666 | 1512121 | |
| 06/05/2020 18:00 | ITA | 15769 | 1333 | 17102 | 74426 | 91528 | -6939 | 1444 | 93245 | 29684 | | | 214457 | 214457 | 2310929 | 1549892 | dc-IT-0021 |
| 07/05/2020 18:00 | ITA | 15174 | 1311 | 16485 | 73139 | 89624 | -1904 | 1401 | 96276 | 29958 | | | 215858 | 215858 | 2381288 | 1563557 | dc-IT-0023 |
| 08/05/2020 18:00 | ITA | 14636 | 1168 | 15804 | 72157 | 87961 | -1663 | 1327 | 99023 | 30201 | | | 217185 | 217185 | 2445063 | 1608985 | dc-IT-0025 |
| 09/05/2020 18:00 | ITA | 13834 | 1034 | 14868 | 69974 | 84842 | -3119 | 1083 | 103031 | 30395 | | | 218268 | 218268 | 2514234 | 1645076 | |
| 10/05/2020 18:00 | ITA | 13618 | 1027 | 14645 | 68679 | 83324 | -1518 | 802 | 105186 | 30560 | | | 219070 | 219070 | 2565912 | 1676460 | |
| 11/05/2020 18:00 | ITA | 13539 | 999 | 14538 | 67950 | 82488 | -836 | 744 | 106587 | 30739 | | | 219814 | 219814 | 2606652 | 1702283 | |
| 12/05/2020 18:00 | ITA | 12865 | 952 | 13817 | 67449 | 81266 | -1222 | 1402 | 109039 | 30911 | | | 221216 | 221216 | 2673655 | 1741903 | dc-IT-0027 |
| 13/05/2020 18:00 | ITA | 12172 | 893 | 13065 | 65392 | 78457 | -2809 | 888 | 112541 | 31106 | | | 222104 | 222104 | 2735628 | 1778952 | |

| | | | | | | | | | | | | | | | | | | |
|---|---|---|---|---|---|---|---|---|---|---|---|---|---|---|---|---|---|---|
| 14/05/2020 18:00 | ITA | | 11453 | 855 | 12308 | 64132 | 76440 | -2017 | 992 | 115288 | 31368 | | | 223096 | 223096 | 2807504 | 1820083 | dc-IT-0029 |
| 15/05/2020 18:00 | ITA | | 10792 | 808 | 11600 | 60470 | 72070 | -4370 | 789 | 120205 | 31610 | | | 223885 | 223885 | 2875680 | 1859110 | dc-IT-0031 |
| 16/05/2020 18:00 | ITA | | 10400 | 775 | 11175 | 59012 | 70187 | -1883 | 875 | 122810 | 31763 | | | 224760 | 224760 | 2944859 | 1899767 | |
| 18/05/2020 18:00 | ITA | | 10311 | 762 | 11073 | 57278 | 68351 | -1836 | 675 | 125176 | 31908 | | | 225435 | 225435 | 3004960 | 1933272 | |
| 18/05/2020 18:00 | ITA | | 10207 | 749 | 10956 | 55597 | 66553 | -1798 | 451 | 127326 | 32007 | | | 225886 | 225886 | 3041366 | 1959373 | pd-IT-0019 |
| 19/05/2020 18:00 | ITA | | 9991 | 716 | 10707 | 54422 | 65129 | -1424 | 813 | 129401 | 32169 | | | 226699 | 226699 | 3104524 | 1999599 | dc-IT-0033 |
| 20/05/2020 18:00 | ITA | | 9624 | 676 | 10300 | 52452 | 62752 | -2377 | 665 | 132282 | 32330 | | | 227364 | 227364 | 3171719 | 2038216 | |
| 21/05/2020 18:00 | ITA | | 9269 | 640 | 9909 | 51051 | 60960 | -1792 | 642 | 134560 | 32486 | | | 228006 | 228006 | 3243398 | 2078860 | |
| 22/05/2020 18:00 | ITA | | 8957 | 595 | 9552 | 49770 | 59322 | -1638 | 652 | 136720 | 32616 | | | 228658 | 228658 | 3318778 | 2121847 | |
| 23/05/2020 18:00 | ITA | | 8695 | 572 | 9267 | 48485 | 57752 | -1570 | 669 | 138840 | 32735 | | | 229327 | 229327 | 3391188 | 2164426 | |
| 24/05/2020 18:00 | ITA | | 8613 | 553 | 9166 | 47428 | 56594 | -1158 | 531 | 140479 | 32785 | | | 229858 | 229858 | 3447012 | 2198632 | pd-IT-0021 |
| 25/05/2020 18:00 | ITA | | 8185 | 541 | 8726 | 46574 | 55300 | -1294 | 300 | 141981 | 32877 | | | 230158 | 230158 | 3482253 | 2219308 | dc-IT-0035 |
| 26/05/2020 18:00 | ITA | | 7917 | 521 | 8438 | 44504 | 52942 | -2358 | 397 | 144658 | 32955 | | | 230555 | 230555 | 3539927 | 2253252 | |
| 27/05/2020 18:00 | ITA | | 7729 | 505 | 8234 | 42732 | 50966 | -1976 | 584 | 147101 | 33072 | | | 231139 | 231139 | 3607251 | 2290551 | |
| 28/05/2020 18:00 | ITA | | 7379 | 489 | 7868 | 40118 | 47986 | -2980 | 593 | 150604 | 33142 | | | 231732 | 231732 | 3683144 | 2330389 | |
| 29/05/2020 18:00 | ITA | | 7094 | 475 | 7569 | 38606 | 46175 | -1811 | 516 | 152844 | 33229 | | | 232248 | 232248 | 3755279 | 2368622 | dc-IT-0037 |
| 30/05/2020 18:00 | ITA | | 6680 | 450 | 7130 | 36561 | 43691 | -2484 | 416 | 155633 | 33340 | | | 232664 | 232664 | 3824621 | 2404673 | nd-IT-0016 |
| 31/05/2020 18:00 | ITA | | 6387 | 435 | 6822 | 35275 | 42097 | -1594 | 355 | 157507 | 33415 | | | 233019 | 233019 | 3878739 | 2433621 | pd-IT-0023 |
| 01/06/2020 18:00 | ITA | | 6099 | 424 | 6523 | 34844 | 41367 | -730 | 178 | 158355 | 33475 | | | 233197 | 233197 | 3910133 | 2451674 | |
| 02/06/2020 18:00 | ITA | | 5916 | 408 | 6324 | 33569 | 39893 | -1474 | 318 | 160092 | 33530 | | | 233515 | 233515 | 3962292 | 2477302 | |
| 03/06/2020 18:00 | ITA | | 5742 | 353 | 6095 | 33202 | 39297 | -596 | 321 | 160938 | 33601 | | | 233836 | 233836 | 3999591 | 2497337 | |
| 04/06/2020 18:00 | ITA | | 5503 | 338 | 5841 | 32588 | 38429 | -868 | 177 | 161895 | 33689 | | | 234013 | 234013 | 4049544 | 2524788 | dc-IT-0039 |
| 05/06/2020 18:00 | ITA | | 5301 | 316 | 5617 | 31359 | 36976 | -1453 | 518 | 163781 | 33774 | | | 234531 | 234531 | 4114572 | 2565258 | |
| 06/06/2020 18:00 | ITA | | 5002 | 293 | 5295 | 30582 | 35877 | -1099 | 270 | 165078 | 33846 | | | 234801 | 234801 | 4187057 | 2599294 | |
| 07/06/2020 18:00 | ITA | | 4864 | 287 | 5151 | 30111 | 35262 | -615 | 197 | 165837 | 33899 | | | 234998 | 234998 | 4236535 | 2627188 | |
| 08/06/2020 18:00 | ITA | | 4729 | 283 | 5012 | 29718 | 34730 | -532 | 280 | 166584 | 33964 | | | 235278 | 235278 | 4263647 | 2643489 | |
| 09/06/2020 18:00 | ITA | | 4581 | 263 | 4844 | 28028 | 32872 | -1858 | 283 | 168646 | 34043 | | | 235561 | 235561 | 4318650 | 2675689 | dc-IT-0041;dc-IT-0043 |
| 10/06/2020 18:00 | ITA | | 4320 | 249 | 4569 | 27141 | 31710 | -1162 | 202 | 169939 | 34114 | | | 235763 | 235763 | 4381349 | 2713554 | |
| 11/06/2020 18:00 | ITA | | 4131 | 236 | 4367 | 26270 | 30637 | -1073 | 379 | 171338 | 34167 | | | 236142 | 236142 | 4443821 | 2746545 | |
| 12/06/2020 18:00 | ITA | | 3893 | 227 | 4120 | 24877 | 28997 | -1640 | 163 | 173085 | 34223 | | | 236305 | 236305 | 4514441 | 2784196 | dc-IT-0045;dc-IT-0047 |
| 13/06/2020 18:00 | ITA | | 3747 | 220 | 3967 | 23518 | 27485 | -1512 | 346 | 174865 | 34301 | | | 236651 | 236651 | 4564191 | 2817076 | dc-IT-0049 |
| 14/06/2020 18:00 | ITA | | 3594 | 209 | 3803 | 22471 | 26274 | -1211 | 338 | 176370 | 34345 | | | 236989 | 236989 | 4620718 | 2846621 | |
| 15/06/2020 18:00 | ITA | | 3489 | 207 | 3696 | 22213 | 25909 | -365 | 303 | 177010 | 34371 | | | 237292 | 237290 | 4648825 | 2864084 | dc-IT-0051 |
| 16/06/2020 18:00 | ITA | | 3301 | 177 | 3478 | 21091 | 24569 | -1340 | 210 | 178526 | 34405 | | | 237502 | 237500 | 4695707 | 2891846 | |
| 18/06/2020 18:00 | ITA | | 3113 | 163 | 3276 | 20649 | 23925 | -644 | 329 | 179455 | 34448 | | | 237831 | 237828 | 4773408 | 2925803 | dc-IT-0053 |
| 18/06/2020 18:00 | ITA | | 2867 | 168 | 3035 | 20066 | 23101 | -824 | 333 | 180544 | 34514 | | | 238164 | 238159 | 4831562 | 2958724 | dc-IT-0055 |
| 19/06/2020 18:00 | ITA | | 2632 | 161 | 2793 | 18750 | 21543 | -1558 | 251 | 181907 | 34561 | | | 238415 | 238011 | 4889103 | 2987294 | dc-IT-0057;dc-IT-0059 |
| 20/06/2020 18:00 | ITA | | 2474 | 152 | 2626 | 18586 | 21212 | -331 | 262 | 182453 | 34610 | | | 238677 | 238275 | 4943825 | 3017169 | dc-IT-0061;dc-IT-0063 |
| 21/06/2020 18:00 | ITA | | 2314 | 148 | 2462 | 18510 | 20972 | -240 | 224 | 182893 | 34634 | | | 238901 | 238499 | 4984370 | 3041750 | |
| 22/06/2020 18:00 | ITA | | 2038 | 127 | 2165 | 18472 | 20637 | -335 | 218 | 183426 | 34657 | | | 239119 | 238720 | 5013342 | 3057902 | dc-IT-0065 |
| 23/06/2020 18:00 | ITA | | 1853 | 115 | 1968 | 17605 | 19573 | -1064 | 122 | 184585 | 34675 | | | 239241 | 238833 | 5053827 | 3081127 | dc-IT-0067;dc-IT-0069 |
| 24/06/2020 18:00 | ITA | | 1610 | 107 | 1717 | 16938 | 18655 | -918 | 190 | 186111 | 34644 | | | 239431 | 239410 | 5107093 | 3111364 | dc-IT-0071 |
| 25/06/2020 18:00 | ITA | | 1515 | 103 | 1618 | 16685 | 18303 | -352 | 296 | 186725 | 34678 | 223905 | 15801 | 239727 | 239706 | 5163154 | 3140785 | |
| 26/06/2020 18:00 | ITA | | 1356 | 105 | 1461 | 16177 | 17638 | -665 | 259 | 187615 | 34708 | 223440 | 16521 | 239986 | 239961 | 5215922 | 3169116 | |
| 27/06/2020 18:00 | ITA | | 1260 | 97 | 1357 | 15479 | 16836 | -802 | 175 | 188584 | 34716 | 223262 | 16874 | 240161 | 240136 | 5277273 | 3198837 | |
| 28/06/2020 18:00 | ITA | | 1160 | 98 | 1258 | 15423 | 16681 | -155 | 174 | 188891 | 34738 | 223272 | 17038 | 240335 | 240310 | 5314619 | 3220020 | |
| 29/06/2020 18:00 | ITA | | 1120 | 96 | 1216 | 15280 | 16496 | -185 | 126 | 189196 | 34744 | 223254 | 17182 | 240461 | 240436 | 5341837 | 3235504 | |
| 30/06/2020 18:00 | ITA | | 1090 | 93 | 1183 | 14380 | 15563 | -933 | 142 | 190248 | 34767 | 222901 | 17677 | 240603 | 240578 | 5390110 | 3263975 | |
| 01/07/2020 18:00 | ITA | | 1025 | 87 | 1112 | 14143 | 15255 | -308 | 187 | 190717 | 34788 | 222564 | 18196 | 240790 | 240760 | 5445476 | 3293300 | |
| 02/07/2020 18:00 | ITA | | 963 | 82 | 1045 | 14015 | 15060 | -195 | 201 | 191083 | 34818 | 222464 | 18497 | 240991 | 240961 | 5498719 | 3322447 | |
| 03/07/2020 18:00 | ITA | | 956 | 79 | 1035 | 13849 | 14884 | -176 | 223 | 191467 | 34833 | 222430 | 18754 | 241214 | 241184 | 5548815 | 3348127 | |
| 04/07/2020 18:00 | ITA | | 940 | 71 | 1011 | 13610 | 14621 | -263 | 235 | 191944 | 34854 | 222324 | 19095 | 241449 | 241419 | 5600826 | 3377073 | |
| 05/07/2020 18:00 | ITA | C | | 74 | 1019 | 13623 | 14642 | 21 | 192 | 192108 | 34861 | 222368 | 19243 | 241641 | 241611 | 5638288 | 3398239 | |
| 06/07/2020 18:00 | ITA | | 946 | 72 | 1018 | 13691 | 14709 | 67 | 208 | 192241 | 34869 | 222410 | 19409 | 241849 | 241819 | 5660454 | 3412010 | |
| 07/07/2020 18:00 | ITA | | 940 | 70 | 1010 | 13232 | 14242 | -467 | 138 | 192815 | 34899 | 222292 | 19664 | 241987 | 241956 | 5703673 | 3434500 | |
| 08/07/2020 18:00 | ITA | | 899 | 71 | 970 | 12625 | 13595 | -647 | 193 | 193640 | 34914 | 222145 | 20004 | 242180 | 242149 | 5754116 | 3463179 | |
| 09/07/2020 18:00 | ITA | | 871 | 69 | 940 | 12519 | 13459 | -136 | 229 | 193978 | 34926 | 222106 | 20257 | 242409 | 242363 | 5806668 | 3493126 | |
| 10/07/2020 18:00 | ITA | | 844 | 65 | 909 | 12519 | 13428 | -31 | 276 | 194273 | 34938 | 222079 | 20560 | 242685 | 242639 | 5854621 | 3520377 | |
| 11/07/2020 18:00 | ITA | | 826 | 67 | 893 | 12410 | 13303 | -125 | 188 | 194579 | 34945 | 221994 | 20833 | 242873 | 242827 | 5900552 | 3545826 | |
| 12/07/2020 18:00 | ITA | | 776 | 68 | 844 | 12335 | 13179 | -124 | 234 | 194928 | 34954 | 222036 | 21025 | 243107 | 243061 | 5938811 | 3568887 | |
| 13/07/2020 18:00 | ITA | | 768 | 65 | 833 | 12324 | 13157 | -22 | 169 | 195106 | 34967 | 222074 | 21156 | 243276 | 243230 | 5962744 | 3582893 | |
| 14/07/2020 18:00 | ITA | | 777 | 60 | 837 | 12082 | 12919 | -238 | 114 | 195441 | 34984 | 222053 | 21291 | 243390 | 243344 | 6004611 | 3607115 | |
| 15/07/2020 18:00 | ITA | | 797 | 57 | 854 | 11639 | 12493 | -426 | 163 | 196016 | 34997 | 222075 | 21431 | 243553 | 243506 | 6053060 | 3635507 | |
| 16/07/2020 18:00 | ITA | | 750 | 53 | 803 | 11670 | 12473 | -20 | 230 | 196246 | 35017 | 222079 | 21657 | 243783 | 243736 | 6103492 | 3663596 | |
| 17/07/2020 18:00 | ITA | | 771 | 50 | 821 | 11635 | 12456 | -17 | 233 | 196483 | 35028 | 222072 | 21895 | 244016 | 243967 | 6154259 | 3692257 | |
| 18/07/2020 18:00 | ITA | | 757 | 50 | 807 | 11561 | 12368 | -88 | 249 | 196806 | 35042 | 222138 | 22078 | 244265 | 244216 | 6202524 | 3719826 | |
| 19/07/2020 18:00 | ITA | | 743 | 49 | 792 | 11648 | 12440 | 72 | 219 | 196949 | 35045 | 222180 | 22254 | 244484 | 244434 | 6238049 | 3740447 | |
| 20/07/2020 18:00 | ITA | | 745 | 47 | 792 | 11612 | 12404 | -36 | 190 | 197162 | 35058 | 222212 | 22412 | 244674 | 244624 | 6262302 | 3754568 | |
| 21/07/2020 18:00 | ITA | | 732 | 49 | 781 | 11467 | 12248 | -156 | 129 | 197431 | 35073 | 222232 | 22520 | 244803 | 244752 | 6305412 | 3778483 | |
| 22/07/2020 18:00 | ITA | | 724 | 48 | 772 | 11550 | 12322 | 74 | 282 | 197628 | 35082 | 222220 | 22812 | 245085 | 245032 | 6354730 | 3807771 | |
| 23/07/2020 18:00 | ITA | | 713 | 49 | 762 | 11642 | 12404 | 82 | 306 | 197842 | 35092 | 222279 | 23059 | 245391 | 245338 | 6415041 | 3840789 | |
| 24/07/2020 18:00 | ITA | | 713 | 46 | 759 | 11542 | 12301 | -103 | 252 | 198192 | 35097 | 222294 | 23296 | 245643 | 245590 | 6468375 | 3869759 | |
| 25/07/2020 18:00 | ITA | | 731 | 41 | 772 | 11670 | 12442 | 141 | 275 | 198320 | 35102 | 222348 | 23516 | 245918 | 245864 | 6520046 | 3897818 | |
| 26/07/2020 18:00 | ITA | | 735 | 44 | 779 | 11786 | 12565 | 123 | 255 | 198446 | 35107 | 222382 | 23736 | 246173 | 246118 | 6560572 | 3922995 | |
| 27/07/2020 18:00 | ITA | | 740 | 45 | 785 | 11796 | 12581 | 16 | 170 | 198593 | 35112 | 222442 | 23844 | 246343 | 246286 | 6586123 | 3942369 | |
| 28/07/2020 18:00 | ITA | | 749 | 40 | 789 | 11820 | 12609 | 28 | 212 | 198756 | 35123 | 222473 | 24015 | 246555 | 246488 | 6634293 | 3967710 | |
| 29/07/2020 18:00 | ITA | | 731 | 38 | 769 | 11847 | 12616 | 7 | 289 | 199031 | 35129 | 222569 | 24207 | 246844 | 246776 | 6690311 | 3998585 | |
| 30/07/2020 18:00 | ITA | | 748 | 47 | 795 | 11435 | 12230 | -386 | 386 | 199796 | 35132 | 222720 | 24438 | 247230 | 247158 | 6752169 | 4031981 | |
| 31/07/2020 18:00 | ITA | | 716 | 41 | 757 | 11665 | 12422 | 192 | 379 | 199974 | 35141 | 222744 | 24793 | 247609 | 247537 | 6813113 | 4061667 | |
| 01/08/2020 18:00 | ITA | | 705 | 43 | 748 | 11709 | 12457 | 35 | 295 | 200229 | 35146 | 222815 | 25017 | 247904 | 247832 | 6873496 | 4093572 | |
| 02/08/2020 18:00 | ITA | | 708 | 42 | 750 | 11706 | 12456 | -1 | 239 | 200460 | 35154 | 222877 | 25193 | 248143 | 248070 | 6916765 | 4118068 | |
| 03/08/2020 18:00 | ITA | | 734 | 41 | 775 | 11699 | 12474 | 18 | 159 | 200589 | 35166 | 222891 | 25338 | 248302 | 248229 | 6940801 | 4131535 | |

| | | | | | | | | | | | | | | | | |
|---|---|---|---|---|---|---|---|---|---|---|---|---|---|---|---|---|
| 04/08/2020 18:00 | ITA | 761 | 41 | 802 | 11680 | 12482 | 8 | 190 | 200766 | 35171 | 222954 | 25465 | 248492 | 248419 | 6984589 | 4155026 |
| 05/08/2020 18:00 | ITA | 764 | 41 | 805 | 11841 | 12646 | 164 | 384 | 200976 | 35181 | 223044 | 25759 | 248876 | 248803 | 7041040 | 4184765 |
| 06/08/2020 18:00 | ITA | 762 | 42 | 804 | 11890 | 12694 | 48 | 402 | 201323 | 35187 | 223113 | 26091 | 249278 | 249204 | 7099713 | 4216934 |
| 07/08/2020 18:00 | ITA | 779 | 42 | 821 | 12103 | 12924 | 230 | 552 | 201642 | 35190 | 223201 | 26555 | 249830 | 249756 | 7158909 | 4247326 |
| 08/08/2020 18:00 | ITA | 771 | 43 | 814 | 12139 | 12953 | 29 | 347 | 201947 | 35203 | 223232 | 26871 | 250177 | 250103 | 7212207 | 4273957 |
| 09/08/2020 18:00 | ITA | 763 | 45 | 808 | 12455 | 13263 | 310 | 463 | 202098 | 35205 | 223440 | 27126 | 250640 | 250566 | 7249844 | 4296730 |
| 10/08/2020 18:00 | ITA | 779 | 46 | 825 | 12543 | 13368 | 105 | 259 | 202248 | 35209 | 223556 | 27269 | 250899 | 250825 | 7276276 | 4307634 |
| 11/08/2020 18:00 | ITA | 801 | 49 | 850 | 12711 | 13561 | 193 | 412 | 202461 | 35215 | 223674 | 27563 | 251311 | 251237 | 7316918 | 4329697 |
| 12/08/2020 18:00 | ITA | 779 | 53 | 832 | 12959 | 13791 | 230 | 481 | 202697 | 35225 | 223844 | 27869 | 251792 | 251713 | 7369576 | 4357027 |
| 13/08/2020 18:00 | ITA | 786 | 55 | 841 | 13240 | 14081 | 290 | 523 | 202923 | 35231 | 224058 | 28177 | 252315 | 252235 | 7420764 | 4382656 |
| 14/08/2020 18:00 | ITA | 771 | 56 | 827 | 13422 | 14249 | 168 | 574 | 203326 | 35234 | 224319 | 28490 | 252889 | 252809 | 7467487 | 4407524 |
| 15/08/2020 18:00 | ITA | 764 | 55 | 819 | 13587 | 14406 | 157 | 629 | 203640 | 35392 | 224521 | 28917 | 253518 | 253438 | 7520610 | 4433461 |
| 16/08/2020 18:00 | ITA | 787 | 56 | 843 | 13890 | 14733 | 327 | 479 | 203786 | 35396 | 224694 | 29221 | 253997 | 253915 | 7557417 | 4455931 |
| 17/08/2020 18:00 | ITA | 810 | 58 | 868 | 13999 | 14867 | 134 | 320 | 203968 | 35400 | 224812 | 29423 | 254317 | 254235 | 7588083 | 4477310 |
| 18/08/2020 18:00 | ITA | 843 | 58 | 901 | 14188 | 15089 | 222 | 403 | 204142 | 35405 | 224974 | 29662 | 254720 | 254636 | 7642059 | 4509997 |
| 19/08/2020 18:00 | ITA | 866 | 66 | 932 | 14428 | 15360 | 271 | 642 | 204506 | 35412 | 225215 | 30063 | 255362 | 255278 | 7713154 | 4551287 |
| 20/08/2020 18:00 | ITA | 883 | 68 | 951 | 15063 | 16014 | 654 | 845 | 204686 | 35418 | 225516 | 30602 | 256207 | 256118 | 7790596 | 4600949 |
| 21/08/2020 18:00 | ITA | 919 | 69 | 988 | 15690 | 16678 | 664 | 947 | 204960 | 35427 | 225880 | 31185 | 257154 | 257065 | 7862592 | 4645892 |
| 22/08/2020 18:00 | ITA | 924 | 64 | 988 | 16515 | 17503 | 825 | 1071 | 205203 | 35430 | 226320 | 31816 | 258225 | 258136 | 7940266 | 4692505 |
| 23/08/2020 18:00 | ITA | 971 | 69 | 1040 | 17398 | 18438 | 935 | 1210 | 205470 | 35437 | 226810 | 32535 | 259435 | 259345 | 8007637 | 4739968 |
| 24/08/2020 18:00 | ITA | 1045 | 65 | 1110 | 18085 | 19195 | 757 | 953 | 205662 | 35441 | 227128 | 33170 | 260388 | 260298 | 8053551 | 4773326 |
| 25/08/2020 18:00 | ITA | 1058 | 66 | 1124 | 18590 | 19714 | 519 | 878 | 206015 | 35445 | 227292 | 33882 | 261266 | 261174 | 8125892 | 4819124 |
| 26/08/2020 18:00 | ITA | 1055 | 69 | 1124 | 19629 | 20753 | 1039 | 1367 | 206329 | 35458 | 227723 | 34817 | 262633 | 262540 | 8219421 | 4877178 |
| 27/08/2020 18:00 | ITA | 1131 | 67 | 1198 | 20734 | 21932 | 1179 | 1411 | 206554 | 35463 | 228123 | 35826 | 264044 | 263949 | 8313445 | 4934818 |
| 28/08/2020 18:00 | ITA | 1178 | 74 | 1252 | 21783 | 23035 | 1103 | 1462 | 206902 | 35472 | 228553 | 36856 | 265506 | 265409 | 8410510 | 4999953 |
| 29/08/2020 18:00 | ITA | 1168 | 79 | 1247 | 22909 | 24156 | 1121 | 1444 | 207224 | 35473 | 229002 | 37851 | 266950 | 266853 | 8509618 | 5064247 |
| 30/08/2020 18:00 | ITA | 1251 | 86 | 1337 | 23868 | 25205 | 1049 | 1365 | 207536 | 35477 | 229527 | 38691 | 268315 | 268218 | 8586341 | 5117788 |
| 31/08/2020 18:00 | ITA | 1288 | 94 | 1382 | 24696 | 26078 | 873 | 996 | 207653 | 35483 | 229832 | 39382 | 269311 | 269214 | 8644859 | 5160371 |
| 01/09/2020 18:00 | ITA | 1380 | 107 | 1487 | 25267 | 26754 | 676 | 978 | 207944 | 35491 | 230102 | 40087 | 270289 | 270189 | 8725909 | 5214766 |
| 02/09/2020 18:00 | ITA | 1437 | 109 | 1546 | 26271 | 27817 | 1063 | 1326 | 208201 | 35497 | 230504 | 41011 | 271615 | 271515 | 8828868 | 5280948 |
| 03/09/2020 18:00 | ITA | 1505 | 120 | 1625 | 27290 | 28915 | 1098 | 1397 | 208490 | 35507 | 230950 | 41962 | 273012 | 272912 | 8921658 | 5342150 |
| 04/09/2020 18:00 | ITA | 1607 | 121 | 1728 | 28371 | 30099 | 1184 | 1733 | 209027 | 35518 | 231587 | 43057 | 274745 | 274644 | 9034743 | 5414708 |
| 05/09/2020 18:00 | ITA | 1620 | 121 | 1741 | 29453 | 31194 | 1095 | 1695 | 209610 | 35534 | 232220 | 44118 | 276440 | 276338 | 9142401 | 5484345 |
